# Integrated targeted deep sequencing reveals unique tissue-of-origin and donor-derived cell-free DNA signatures in organ transplant recipients

**DOI:** 10.1101/2025.04.29.25326125

**Authors:** Nicholas Kueng, Fanny Sandberg, Daniel Sidler, Vanessa Banz, Annalisa Berzigotti, Charlotte K. Y. Ng, Carlo R. Largiadèr, Ursula Amstutz

## Abstract

Solid organ transplantation is currently the best option to treat end-stage organ disease. However, it requires lifelong immunosuppressive therapy in most cases, and the diagnosis of rejection and other types of graft injury requires invasive biopsy testing, which poses significant challenges. We developed a targeted deep sequencing assay that extends conventional donor-derived cell-free DNA (dd-cfDNA) analysis by incorporating tissue-of-origin information with the aim of improved non-invasive monitoring of transplant recipients. In this study, plasma cfDNA from liver transplant (LT) and kidney transplant (KT) recipients was analyzed alongside healthy controls to characterize cfDNA release and clearance patterns from graft and recipient cell types, and to identify potential tissue injury signatures. Our assay accurately detected low-abundance, tissue-specific cfDNA, revealing unique cfDNA release patterns associated with the transplanted organ type in recipients with stable allografts. In the early post-transplant period, LT and KT patients exhibited different cfDNA kinetics in numerous tissues, reflecting variation in the response and recovery following reperfusion injury and surgical trauma. Furthermore, the comparison of tissue- and donor-specific cfDNA proportions within 24 hours after transplantation supports a multisource donor-tissue cfDNA release. These findings suggest that incorporating tissue-of-origin information with dd-cfDNA quantification provides important additional insights for evaluating transplant recipient health.

## Introduction

Organ transplantation is often the best treatment for end-stage organ disease. Despite improvements in survival rates and quality of life for patients, it requires life-long immunosuppressive therapy to prevent the rejection of the transplanted organ. Calcineurin inhibitors (CNI) are frequently employed as a basis of immunosuppressive regimens because of their effectiveness in preventing cellular rejection. However, long-term CNI use has been linked to complications such as chronic kidney injury, metabolic disorders, cardiovascular events and an elevated risk of malignancies (1). While managing these risks is critical, accurately monitoring allograft health is equally essential. Invasive tissue biopsy remains the gold standard for diagnosing allograft rejection and distinguishing it from other forms of graft injury, yet it carries inherent risks and places a significant burden on patients and the healthcare system, particularly when used in a surveillance setting.

Donor-derived cell-free DNA (dd-cfDNA) is emerging as a powerful, non-invasive biomarker that reflects tissue injury and cellular damage, making it a promising tool for allograft monitoring (2–4) with the potential to reduce the need for tissue biopsies (5). Current methods for dd-cfDNA quantification, such as next-generation sequencing (NGS) and droplet digital PCR (ddPCR), leverage genetic differences between donor and recipient by analyzing single-nucleotide polymorphisms (SNP) (6,7), indels (8) or mismatches in the *HLA-DRB1* gene (9). However, dd-cfDNA methods have limitations and cannot be applied effectively in all clinical scenarios, such as when the donor and recipient are identical twins or when the recipient has received multiple organs from the same donor. Additionally, some dd-cfDNA may not originate from the transplanted organ itself but from co-transplanted tissue or donor-derived immune cells, particularly in the early post-transplantation period (10).

An alternative approach to detect cfDNA from the transplanted organ, without relying on donor-recipient genetic differences, involves tissue-of-origin-specific DNA methylation patterns. DNA methylation is an epigenetic modification that is distinct for every cell type and is stable and highly reproducible across individuals (11), enabling the identification of cfDNA derived from specific cell types, including the transplanted organ. By examining unique methylation signatures associated with individual cell types, this approach has shown the potential to accurately determine the source of cfDNA (10–13). This strategy could not only be particularly useful in cases where genotype-based dd-cfDNA detection is ineffective but also expand our understanding of cfDNA release in transplant recipients beyond the allograft. However, current methylation-based approaches face technical challenges. Whole-genome bisulfite sequencing (WGBS) provides comprehensive CpG coverage but is too costly for routine clinical use, mainly when performed at higher depths to detect cell types of lower abundance. Widely used targeted methods, such as arrays or reduced-representation bisulfite sequencing, miss key CpG sites informative of the tissue-of-origin (TOO) (11), limiting their utility in such studies. Targeted sequencing offers a balanced solution, enabling a focus on the most informative CpG sites at high sequencing depth to enhance resolution with the possibility of extending to other markers of interest while remaining cost-effective and suitable for high-throughput applications.

While previous studies have investigated the dynamics of dd-cfDNA in transplant recipients (14–16), an important knowledge gap remains regarding the precise origin of cfDNA in the critical early post-transplantation phase. Existing methods distinguishing cfDNA based on donor-recipient genetic differences are unable to identify the specific cellular or tissue sources contributing to cfDNA levels immediately after transplantation, thus possibly not fully leveraging the diagnostic potential. Furthermore, while dd-cfDNA levels in stable transplant recipients have been relatively well-characterized, it is still unclear how the release of cfDNA from a transplanted organ compares to that from healthy individuals or how cfDNA release from recipient tissues, possibly influenced by immunosuppressive therapy or immunologic processes related to the allograft, may be altered in the overall cfDNA pool. Increasing the knowledge of such baseline data can thus improve the use of cfDNA as a marker for transplant health by increasing our understanding of cfDNA release from donor and recipient tissues, which may vary between organ type or recipient condition.

Herein, we describe a newly designed targeted deep sequencing assay leveraging multi-omic information to simultaneously and accurately determine both cfDNA TOO and dd-cfDNA proportions and quantities. We applied this assay in different phases of the solid organ transplant journey, including early after transplantation and through the first year. This approach enabled us to investigate cfDNA release dynamics and to further compare cfDNA profiles between stable transplant recipients and healthy individuals, as well as between kidney and liver recipients.

## Results

### Experimental approach

We developed an experimental workflow to analyze cfDNA in transplant recipients using targeted deep methylation sequencing. In this workflow, cfDNA extracted from plasma underwent enzymatic conversion, followed by single-stranded library preparation and hybridization capture for targeted enrichment using a custom panel. Targeted sequencing was then performed, enabling simultaneous determination of tissue-specific cfDNA methylation patterns, using a panel comprising 975 markers representative of 40 distinct cell types, and quantification of dd-cfDNA, based on 300 SNPs across the human genome. The method was then applied to plasma samples from stable liver (stableLT) and kidney transplant (stableKT) recipients and profiles were compared to those from age- and sex-matched healthy controls (HC, Figure 1). Finally, paired blood samples from liver transplant (LT) and kidney transplant (KT) recipients collected at two early post-transplant time points were examined to characterize the kinetics of cfDNA release and clearance (Figure 1).

**Figure 1:**
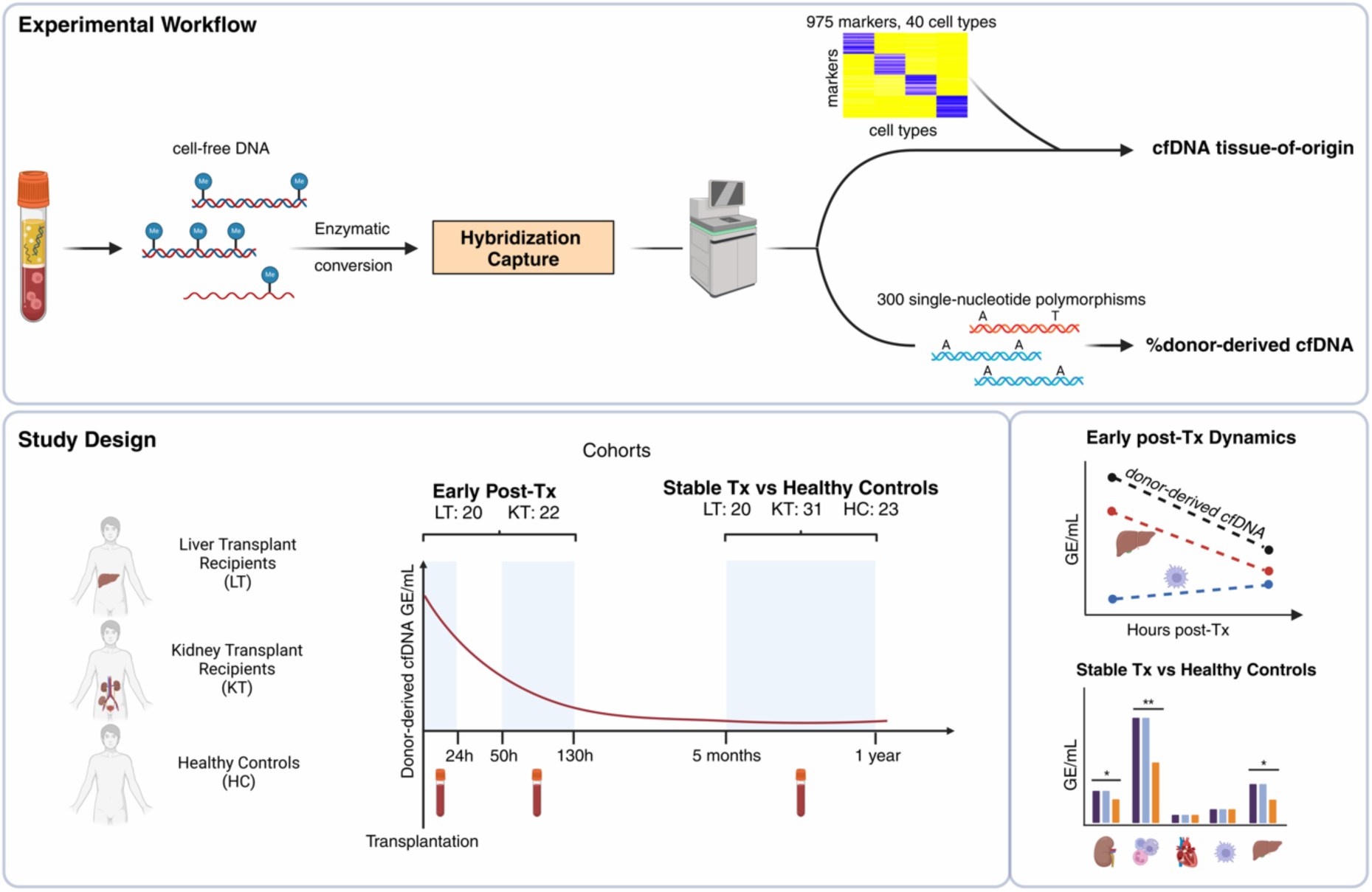
Study overview with experimental workflow and study design. cfDNA was extracted from blood samples. Enzymatically converted cfDNA libraries were enriched for TOO informative markers and SNPs for dd-cfDNA quantification using a novel hybridization capture panel. The bioinformatic analysis resulted in two key metrics: cfDNA TOO after deconvolution based on 975 markers across 40 cell types and dd-cfDNA quantification using 300 SNPs. In the Early post-Tx cohort, blood was collected twice per patient early post-transplantation: the first sample within 24 h and the second between 50 to 130 h after transplantation. In the Stable Tx vs Healthy Controls cohort, blood was collected from healthy control individuals and stable liver and kidney transplant recipients between 5 months and 1 year post-transplantation.

### Validation of targeted sequencing assay for cfDNA tissue-of-origin and dd-cfDNA quantification

To validate our targeted sequencing panel, we analyzed pure primary human cell types relevant for transplant monitoring, including kidney epithelial cells, hepatocytes, B cells and T lymphocytes. TOO analysis of these samples yielded 93 to 98% purity for the expected cell type, demonstrating the specificity of the panel (Figure 2A). Additionally, whole blood from HC (n = 10) was analyzed, revealing that the estimated cfDNA TOO proportions of granulocytes, monocytes and lymphocytes were highly consistent with differential complete blood count (CBC) measurements (Pearson’s r = 0.81–0.99, P < 0.01; Figure 2B). In a subset of 68 transplant recipient samples, we independently quantified the fraction of dd-cfDNA (%dd-cfDNA) using a ddPCR-based method. The %dd-cfDNA determined by our targeted sequencing assay exhibited a strong correlation with quantities obtained by ddPCR (Pearson’s r = 0.99, P < 0.01; Figure 2C). We further assessed the workflow performance by conducting in silico mixture experiments. Reads from kidney epithelial cells or hepatocytes were computationally mixed with white blood cell (WBC) data from three HC (with an additional 3% hepatocytes included in kidney-WBC mixtures for a plasma-like background) at varying proportions and sequencing depths (20 replicates per combination). The workflow accurately detected kidney epithelial and hepatocyte fractions as low as 0.3% (Figure 2D), with the coefficient of variation (CV) decreasing with increasing sequencing depth. CVs below 20% were achieved at a depth of coverage of 400x or higher for spike-in fractions of 0.3% (Supplementary Figure 4). A similar in silico approach was applied to dd-cfDNA quantification using mixtures of plasma cfDNA data from two HC. The workflow reliably detected dd-cfDNA at fractions as low as 0.1% (Figure 2E), with CVs below 20% at ≥ 400x coverage and below 30% at ≥ 300x coverage (Supplementary Figure 5). In HC (n = 23), background levels were low, with a median of 0.097% (interquartile range, IQR: 0.073–0.136%) of non-HC allele signal. Across all plasma samples analyzed in this study (n = 158), the median sequencing coverage was 1’119x (IQR: 913–1’377) for the TOO informative regions and 884x (IQR: 745–1’082) for SNP targets. These depths are approximately threefold and twofold higher, respectively, than the minimum coverage required to achieve a CV equal or smaller than 20% for detecting 0.3% cell type contributions (kidney epithelium and hepatocyte) and 0.1% dd-cfDNA. Lastly, the enzymatic conversion yielded good conversion rates with a median of 99.7% (IQR: 99.6–99.8%), with all samples exceeding 96%. Together, these results underscore the robustness and sensitivity of our workflow for both TOO analysis and dd-cfDNA quantification.

**Figure 2:**
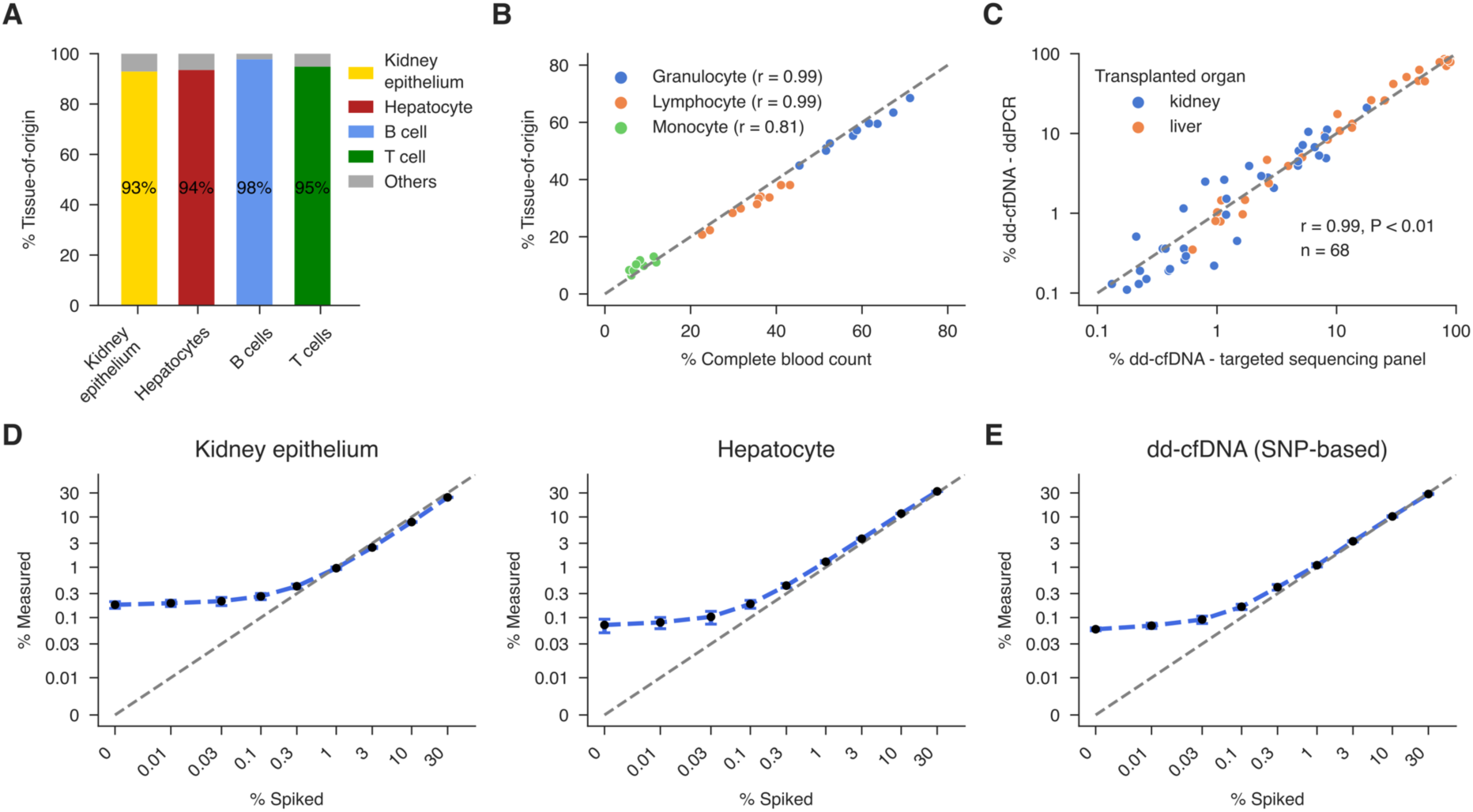
Analytical performance of the targeted sequencing workflow for cfDNA TOO determination and dd-cfDNA quantification. (**A**) Stacked bar chart showing measured TOO of genomic DNA (gDNA) from primary human cell types: kidney epithelium (n = 1), hepatocytes (n = 1), B cells (n = 2) and T cells (n = 1). Percentages in each bar represent the proportion of the measured cell type. All non-target cell type proportions were summed up as “Others”. (**B**) Correlation between measured TOO of whole blood gDNA and the corresponding granulocyte, lymphocyte and monocyte complete blood count proportions from 10 HC. Each data series (colored points) represents one cell type with significant (P < 0.05) Pearson’s correlation coefficients shown in the legend. (**C**) Comparison of dd-cfDNA measurements by ddPCR (y-axis) versus the targeted sequencing panel (x-axis) for kidney (orange) and liver (blue) transplant recipients, demonstrating high concordance (Pearson’s r). (**D**) TOO deconvolution results of in silico mixtures. Reads from sequenced gDNA from kidney epithelium or hepatocyte samples were computationally mixed into a background of leukocyte gDNA reads from three HC. The background for the kidney epithelium mixtures consisted of 97% leukocyte and 3% hepatocyte reads. The mixture shown here had a sequencing depth of 1’000x. Black markers show the median-determined contribution for 20 replicates with the error bars displaying one standard deviation. (**E**) dd-cfDNA proportion results of in silico mixtures. Plasma cfDNA reads from two HC were mixed at various fractions. The mixture shown here had a sequencing depth of 1’000x. Black markers show the median-determined contribution for 20 replicates, with the error bars displaying one standard deviation. The grey line represents the identity line where displayed (y = x).

### TOO profiling reveals distinct cfDNA signatures in stable transplant recipients

TOO deconvolution enables a detailed examination of cfDNA release in transplant recipients and potentially organ-specific effects of immunosuppressive therapy. To determine the cfDNA TOO signatures, total cfDNA was extracted and quantified from plasma of stableLT (n = 20) and stableKT (n = 31) recipients and compared with a sex- and age-matched HC cohort (n = 23, Table 1). Total plasma cfDNA was significantly elevated in transplant recipients (P < 0.001) with 4.5-fold higher levels in stableLT (genome equivalents/mL: GE/mL; median: 7’619 GE/mL, IQR: 3’644–11’370 GE/mL) and 3.5-fold higher levels in stableKT recipients (median: 5’878 GE/mL, IQR: 3’136–8’832 GE/mL) compared to HC (median: 1’627 GE/mL, IQR: 1’300–2’216 GE/mL; Figure 3A) with the majority (>95%; Supplementary Figure 9) of the cfDNA being recipient-derived in both transplant recipient groups. To analyze the source of plasma cfDNA, we performed TOO deconvolution. Hematological-derived cfDNA constituted the largest fraction of total cfDNA in all three groups (Figure 3B), with the relative contributions from different cell types varying considerably between groups, indicating distinct release dynamics in transplant recipients. To assess differences between groups for each cell type, the cell type-specific fractions were multiplied by the total concentration of cfDNA to calculate the absolute GE/mL. The overall higher cfDNA quantities in transplant recipients were predominantly driven by a significant increase in granulocyte-derived cfDNA (P < 0.0001; Figure 3B/C). Notably, transplant recipients also displayed a median of 3- to 5-fold higher levels of non-hematological cfDNA compared with HC (P < 0.001; Supplementary Figure 10). As shown in Figure 3D, comparisons across 40 cell types among the three groups revealed differences in cfDNA quantities between at least two groups for 29 of the 40 cell types. Twelve cell types differed both between stableLT vs. HC and stableLT vs. stableKT, and seven differed between HC and both transplant groups. Only two cell types exhibited distinct differences between the two transplant groups, and for all significant comparisons, HC consistently displayed lower absolute cfDNA levels (Supplementary Figure 11). Moreover, cfDNA from multiple cell types was uniquely detectable only in transplant patients. Among non-hematological cell types, endothelial cells and hepatocytes were the predominant contributors to cfDNA (Figure 3E/F). Given that nephrotoxicity mediated by endothelial injury is a well-recognized complication of CNIs (17), we examined tissue-specific cfDNA alterations for kidney epithelium. Most HC had no detectable cfDNA from the kidney epithelium but unexpectedly the stableLT group showed an increase with levels significantly higher than those in both the HC and the stableKT groups (Figure 3G). Similarly, endothelial-derived cfDNA was markedly increased (2-fold) in both transplant groups (P < 0.001; Figure 3E). In stableLT, hepatocyte-derived cfDNA was significantly higher with a 2-fold increase compared to HC and stableKT (Figure 3F). Interestingly, cfDNA from other major hematological sources, including monocytes/macrophages (P < 0.05; Figure 3H) and erythroid progenitors (P < 0.01; Figure 3I) also showed differences between the groups. In addition, cfDNA from all pancreatic cell types combined was elevated in transplant recipients compared to HC (P < 0.05; Figure 3J) with higher levels in stableLT compared to stableKT. Acinar, beta and delta cell-derived cfDNA was increased exclusively in stableLT compared to HC (Supplementary Figure 11).

**Figure 3:**
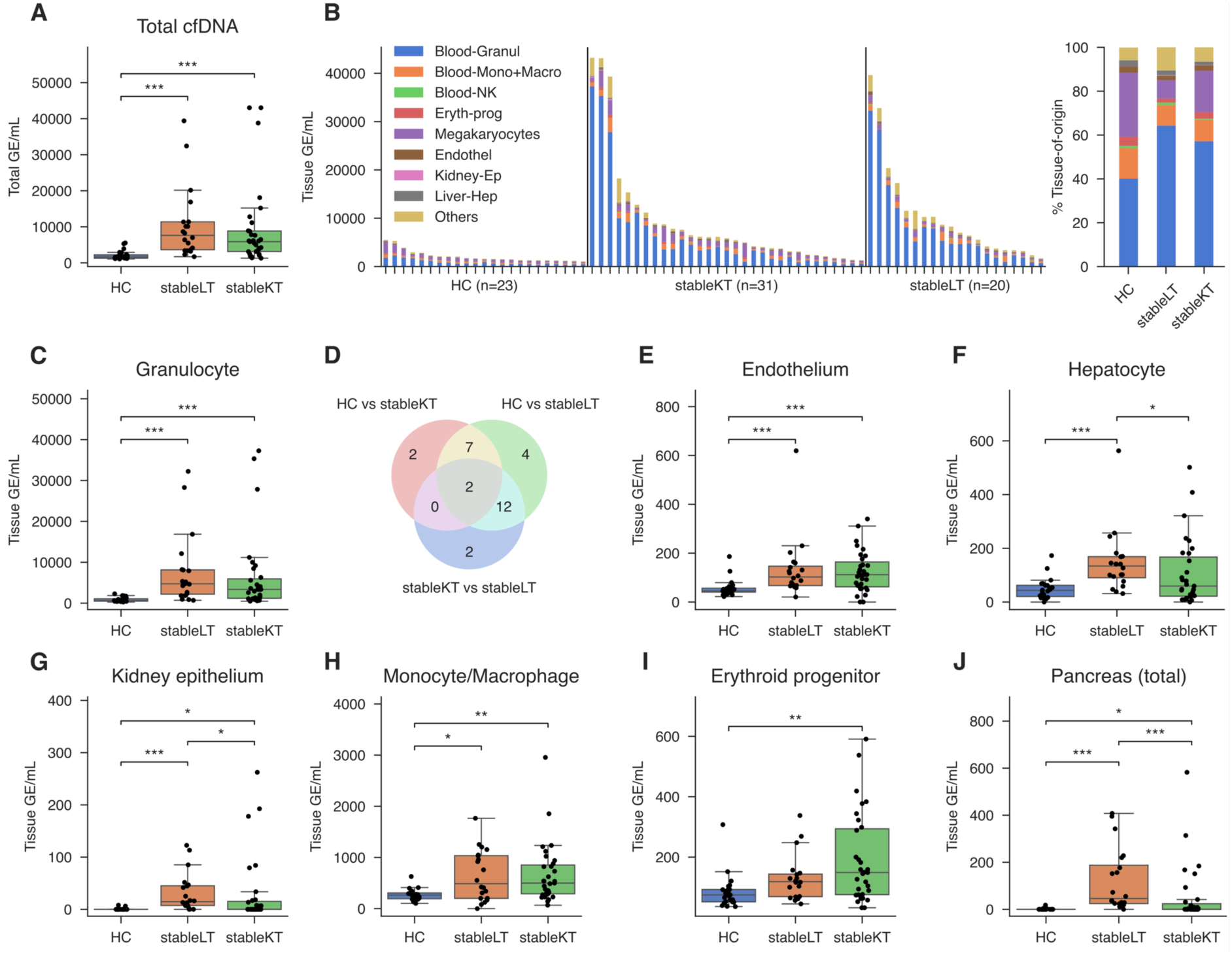
Comparative analysis of tissue-of-origin in healthy controls and stable transplant recipients. (**A**) Total cfDNA. (**B**) Left: Absolute TOO GE/mL in individual samples, with bars grouped by cohort (HC, stableLT, stableKT) in descending order of total GE/mL. Right: Stacked bar chart of mean TOO proportions in HC, stableLT and stableKT; Each colored segment represents a distinct cell type or group. (**C**) Box plots comparing the absolute granulocyte cfDNA quantities across stableLT (n = 20) and stableKT (n = 31) recipient and HC (n = 23). (**E–J**) Box plots comparing the absolute quantities originating from different cell types across the three groups: (**E**) endothelial cells, (**F**) hepatocytes, (**G**) kidney epithelium, (**H**) monocytes and macrophages, (**I**) erythroid progenitors and (**J**) all cell types from the pancreas combined. For each cell type, the Kruskal-Wallis test was performed for global comparisons, with the Benjamini-Hochberg procedure applied to control the false discovery rate (FDR) across cell types. For significant results, Dunn’s post-hoc test was used for pairwise comparisons between groups, and its P values were adjusted using the Bonferroni correction to account for multiple comparisons between groups. * P < 0.05, ** P < 0.01, *** P < 0.001, **** P < 0.0001, no comparison annotation means P > 0.05. The box spans the 25^th^ to 75^th^ percentiles with a line at the median. Whiskers extend to 1.5x the interquartile range.

**Table 1:** Demographic and clinical characteristics of the study population.

| Recipient Characteristics | Stable vs. Healthy Cohort |  |  | Early Post Transplantation Cohort |  |
| --- | --- | --- | --- | --- | --- |
|  | Healthy Controls<br>(n=23) | Stable Liver Recipients<br>(n=20) | Stable Kidney Recipients<br>(n=31) | Early post-TPL Kidney<br>(n=22) | Early post-TPL - Liver<br>(n=20) |
| Sex, n (%) |  |  |  |  |  |
| Male | 18 (78.3%) | 14 (70.0%) | 20 (64.5%) | 13 (59.1%) | 12 (60.0%) |
| Female | 5 (21.7%) | 6 (30.0%) | 11 (35.5%) | 9 (40.9%) | 8 (40.0%) |
| Age (yr), median [IQR] | 54.0 [42.0, 58.0] | 59.0 [47.3, 63.3] | 54.0 [41.5, 65.5] | 58.0 [43.3, 64.8] | 61.0 [54.8, 63.5] |
| Time post-transplantation (days), median [IQR] |  | 111.5 [79.5, 166.5] | 101.0 [87.0, 174.0] |  |  |
| Immunosuppression medication, n (%) |  |  |  |  |  |
| CNI + MPA/MMF + PRED |  | 1 (5.0%) | 29 (93.5%) |  |  |
| CNI + mTORi + MPA/MMF |  | 2 (10.0%) |  |  |  |
| CNI + MPA/MMF |  | 10 (50.0%) |  | n.a. | n.a. |
| CNI + PRED |  | 2 (10.0%) |  |  |  |
| CNI |  | 4 (20.0%) | 1 (3.2%) |  |  |
| mTORi + PRED |  | 1 (5.0%) |  |  |  |
| MPA/MMF + PRED |  |  | 1 (3.2%) |  |  |
| Total ischemia time<sup>a</sup> (min.), median [IQR] |  | 368 [319, 402] | 327 [169, 402] | 325 [185, 424] | 399 [337, 447] |
| Donor Type, n (%) |  |  |  |  |  |
| Living |  | 0 (0.0%) | 14 (45.2%) | 9 (40.9%) | 0 (0.0%) |
| Deceased |  | 20 (100.0%) | 17 (54.8%) | 13 (59.1%) | 20 (100.0%) |
| Delayed graft function, n (%) |  |  |  |  |  |
| yes |  | n.a. | n.a. | 7 (31.8%) | n.a. |
| no |  |  |  | 15 (68.2%) |  |
| Indication for transplantation, n (%) |  |  |  |  |  |
| ADPKD |  |  | 6 (19.4%) | 7 (31.8%) | 1 (5.0%) |
| Diabetes/hypertension/vascular etiology |  |  | 6 (19.4%) | 7 (31.8%) |  |
| Glomerulosclerosis/-nephritis |  |  | 7 (22.6%) | 3 (13.6%) |  |
| Alcohol-related liver disease |  | 3 (15.0%) |  |  | 2 (10.0%) |
| Autoimmune |  | 2 (10.0%) |  |  | 2 (10.0%) |
| Hepatocellular carcinoma |  | 6 (30.0%) |  |  | 6 (30.0%) |
| Metabolic liver disease |  | 3 (15.0%) |  |  | 4 (20.0%) |
| Virus-induced liver cirrhosis |  | 2 (10.0%) |  |  | 2 (10.0%) |
| Other<sup>b</sup> |  | 4 (20.0%) | 12 (38.8%) | 5 (22.7%) | 3 (15.0%) |

post-TPL: post-transplantation, IQR: interquartile range, CNI: calcineurin inhibitors (tacrolimus/cyclosporin A), mTORi: mTOR inhibitor (everolimus), MPA: mycophenolic acid, MMF: mycophenolate mofetil, PRED: prednisolone, ADPKD: autosomal dominant polycystic kidney disease, n.a.: not available. <sup>A</sup> cold and warm ischemia time combined. <sup>B</sup> includes COVID-19, genetic, trauma, birth defect, etc.

We also quantified dd-cfDNA, which is commonly reported as a fraction of the total cfDNA and compared it to the proportion of graft-derived cfDNA determined through TOO deconvolution. In stableLT, the median %dd-cfDNA was 1.4% (IQR: 0.88–2.6%; Supplementary Figure 9), which closely matched the median hepatocyte-derived cfDNA at 1.3% (IQR: 1.1–2.3%; P = 0.41). By contrast, stableKT samples exhibited a median %dd-cfDNA of 0.26% (IQR: 0.22–0.33%; Supplementary Figure 9) and a median kidney epithelium contribution of 0.00% (IQR: 0.00–0.27%), a statistically significant difference (P < 0.001) given that most stableKT samples had kidney epithelium fractions below the estimated detection limit of 0.3%.

### Correlation analysis of cell type-specific cfDNA with clinical parameters in stable transplant recipients

To further elucidate the potential clinical significance of our cfDNA measurements, we assessed the correlations between cell-specific cfDNA levels and established biomarkers of immunosuppression, immune response and organ injury. In a subset of stableKT (n = 20) with available matched cyclosporin A plasma levels, absolute endothelium-derived cfDNA levels correlated moderately with cyclosporin A concentrations (Pearson’s r = 0.54, P < 0.01; Supplementary Figure 12), whereas kidney epithelial cfDNA (P = 0.91) or total cfDNA (P = 0.82) levels showed no correlation. Previous research suggests that immune cfDNA dynamics reflect immune response and turnover rather than changes in the peripheral cell counts (18). Here, matched immune cell concentrations in blood from the stable cohort revealed no correlation of total leukocyte counts with total cfDNA from leukocytes (n = 48; P = 0.94), total lymphocyte counts with total cfDNA from lymphocytes (n = 27; P = 0.60), monocyte counts with monocyte cfDNA (n = 28; P = 0.07) and total neutrophil counts with granulocyte cfDNA (n = 28; P = 0.30). Of note, the median leukocyte count was 5.8 x10^9^/L (IQR: 4.1–8.3 x10^9^/L), with most measurements (81%) within the reference range (3–10.5 x10^9^/L). In a subset of patients with C-reactive protein (CRP) levels measured at the same time point, we did not observe any correlation with total cfDNA (n = 45; P = 0.61). Finally, liver enzyme measurements, which serve as established biomarkers for liver injury, were obtained for stableLT and stableKT recipients where available from the clinical records. Hepatocyte cfDNA showed a moderate correlation with gamma-glutamyl transferase (n = 35; Pearson’s r = 0.54, P < 0.01), alanine aminotransferase (ALAT; n = 39; Pearson’s r = 0.40, P < 0.05) and aspartate aminotransferase (ASAT; n = 33; Pearson’s r = 0.49, P < 0.01) (Supplementary Figure 13).

### Organ-specific dynamics of cfDNA release in the early post-transplant period

Early kinetics of dd-cfDNA have been shown to predict graft recovery and long-term function (19) in KT recipients. However, the precise cellular origins of cfDNA during this critical early phase remain largely undefined. To address this, we evaluated the kinetics of cfDNA release from different cell types in LT and KT recipients by collecting blood samples at two time points. Blood samples were obtained from 20 LT and 22 KT recipients (Table 1) within 24 hours post-transplant (median: 15 h, IQR: 12– 20 h) and between 50–130 hours post-transplantation (median: 69 h, IQR: 62–91 h). These intervals capture key phases of surgical recovery, including responses to trauma, ischemia-reperfusion injury and tissue repair. In LT recipients, total cfDNA levels were substantially elevated in the early post-transplantation period (median: 456’936 GE/mL, IQR: 143’150–693’807 GE/mL) and decreased sharply by 50–130 h (median: 27’396 GE/mL, IQR: 14’105–59’293 GE/mL; P < 0.001; Supplementary Figure 14B). Conversely, KT recipients exhibited lower initial cfDNA concentrations (median: 6’050 GE/mL, IQR: 4’357–12’845 GE/mL) that increased significantly at the later time point (median: 17’786 GE/mL, IQR: 12’953–32’374 GE/mL; P < 0.01; Supplementary Figure 14A). TOO deconvolution in LT recipients revealed that cfDNA from 22 of 40 cell types was significantly elevated in the early phase compared to the second time point (Supplementary Figure 16). Interestingly, kidney epithelial cfDNA levels in KT recipients remained consistently low (P = 0.60; Figure 4A) for most patients. Conversely, LT recipients showed an almost 55-fold decrease in hepatocyte-derived cfDNA from the early to later phase (P < 0.001; Figure 4A), with granulocyte- and endothelium-derived cfDNA also decreasing (P < 0.05; Figures 4B/C). In KT recipients, significant changes between time points were primarily confined to granulocyte-derived cfDNA, which increased at 50–130 h (Figure 4B). While skeletal muscle cfDNA was barely detectable in HC and KT early post-transplantation, LT recipients displayed significantly elevated skeletal muscle cfDNA levels within the first 24 h (median: 9’542 GE/mL, IQR: 645–27’876 GE/mL), which dropped markedly by 50–130 h post-transplantation (median: 132 GE/mL, IQR: 0–280 GE/mL; P < 0.01; Figure 4D). In KT recipients, neither endothelial nor skeletal muscle cfDNA levels differed significantly between the two time points (P > 0.05; Figures 4C/D).

**Figure 4:**
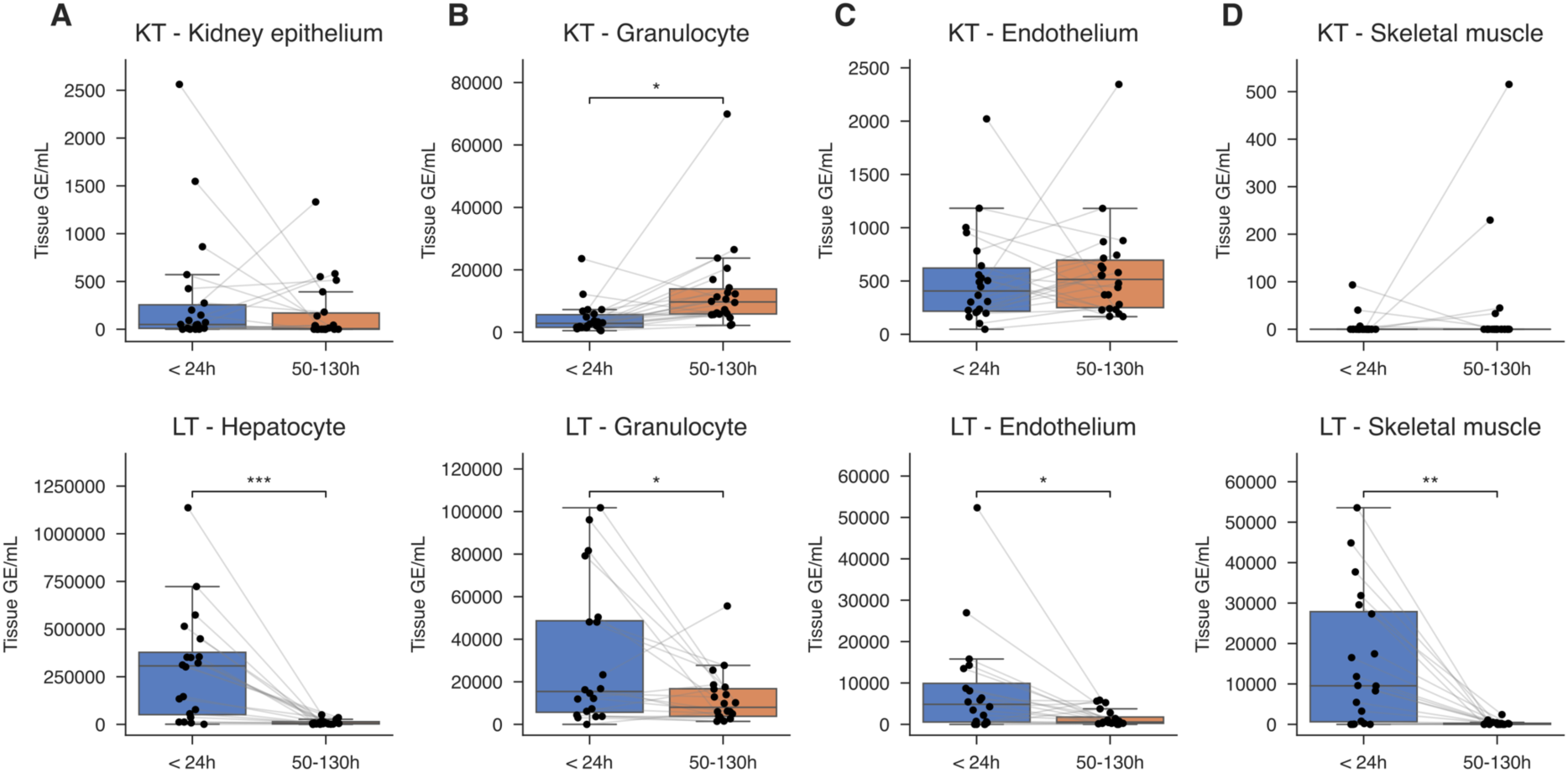
Differences in cell type-specific cfDNA release for kidney and liver transplant recipients early post-transplantation. Cell type-specific cfDNA quantities in KT and LT recipients were measured at < 24 h and 50–130 h post-transplantation. Shown are the cfDNA quantities from (**A**) the kidney epithelium in KT and hepatocytes in LT, (**B**) granulocytes from both, (**C**) endothelium from both and (**D**) skeletal muscle from both, with grey lines connecting repeated measurements from the same individual. The Wilcoxon signed-rank test was used, and P values were adjusted for multiple comparisons across cell types using the Benjamini-Hochberg procedure to control the false discovery rate (FDR). * P < 0.05, ** P < 0.01, *** P < 0.001, **** P < 0.0001, no comparison annotation means P > 0.05. The box spans the 25th to 75th percentiles with a line at the median. Whiskers extend to 1.5x the interquartile range.

Similarly to the stable cohort, blood leukocyte cell counts were routinely measured in this cohort. However, we did not observe any significant correlation between matched total leukocyte counts and total leukocyte-derived cfDNA (n = 83; P = 0.64). This lack of correlation was consistent across each time point and transplanted organ (all correlations: P > 0.05). We also obtained liver enzyme measurements for this cohort, which were only available for LT recipients. We performed correlation analysis between these measurements and absolute hepatocyte cfDNA quantities. The analysis revealed significant correlations with ALAT within 24 h (n = 18; Pearson’s r = 0.57, P < 0.05) and 50–130 h post-transplantation (n = 20; Pearson’s r = 0.56, P < 0.01) (Supplementary Figure 17). The findings were similar for ASAT within 24 h post-transplantation (n = 18; Pearson’s r = 0.80, P < 0.001) but the correlation was not significant 50–130 h post-transplantation (n = 19; P = 0.07) (Supplementary Figure 17).

### Comparison of donor-derived and tissue-specific dynamics in post-transplant plasma

Given that prior studies have predominantly focused on dd-cfDNA as a biomarker for allograft injury, the specific contribution of the primary transplanted tissue to the plasma cfDNA pool remains unclear. To elucidate this relationship, we quantified both the relative and absolute levels of dd-cfDNA in post-transplant samples and compared these values to the graft-derived cfDNA determined by TOO deconvolution. In LT recipients, dd-cfDNA constituted 80.9% (IQR: 71.7–89.9%) of the total cfDNA in the early post-transplant period, decreasing to 29.5% (IQR: 17.6–40.9%) at the later time point. Similarly, absolute dd-cfDNA levels declined from 387’650 GE/mL (IQR: 76’238–562’589 GE/mL) to 7’666 GE/mL (IQR: 3’117–19’255 GE/mL; P < 0.001). The relationship between %dd-cfDNA and the hepatocyte-derived cfDNA proportion within 24 h post-transplantation is characterized by higher dd-cfDNA compared to hepatocyte-derived cfDNA in all patients (Figure 5A), with a median %dd-cfDNA not explained by hepatocyte-derived cfDNA of 17.3% (IQR: 8.0–25.2%). We hypothesized that, in addition to hepatocytes, donor-derived endothelial cells and granulocytes might contribute to the dd-cfDNA. When the proportions of cfDNA from endothelium and granulocytes were added to that from hepatocytes, we observed a strong correlation with the overall %dd-cfDNA (Pearson’s r = 0.89; Figure 5A). In KT recipients, the percentage of dd-cfDNA decreased significantly from 3.9% (IQR: 1.6–7.0%) in samples collected within 24 h post-transplant to 0.53% (IQR: 0.38–1.7%) at 50–130 h (P < 0.001). In contrast, the absolute dd-cfDNA did not differ significantly between the two time points (median: 322 GE/mL, IQR: 140–847 GE/mL vs. median: 112 GE/mL, IQR: 57–360 GE/mL; P = 0.08).

**Figure 5:**
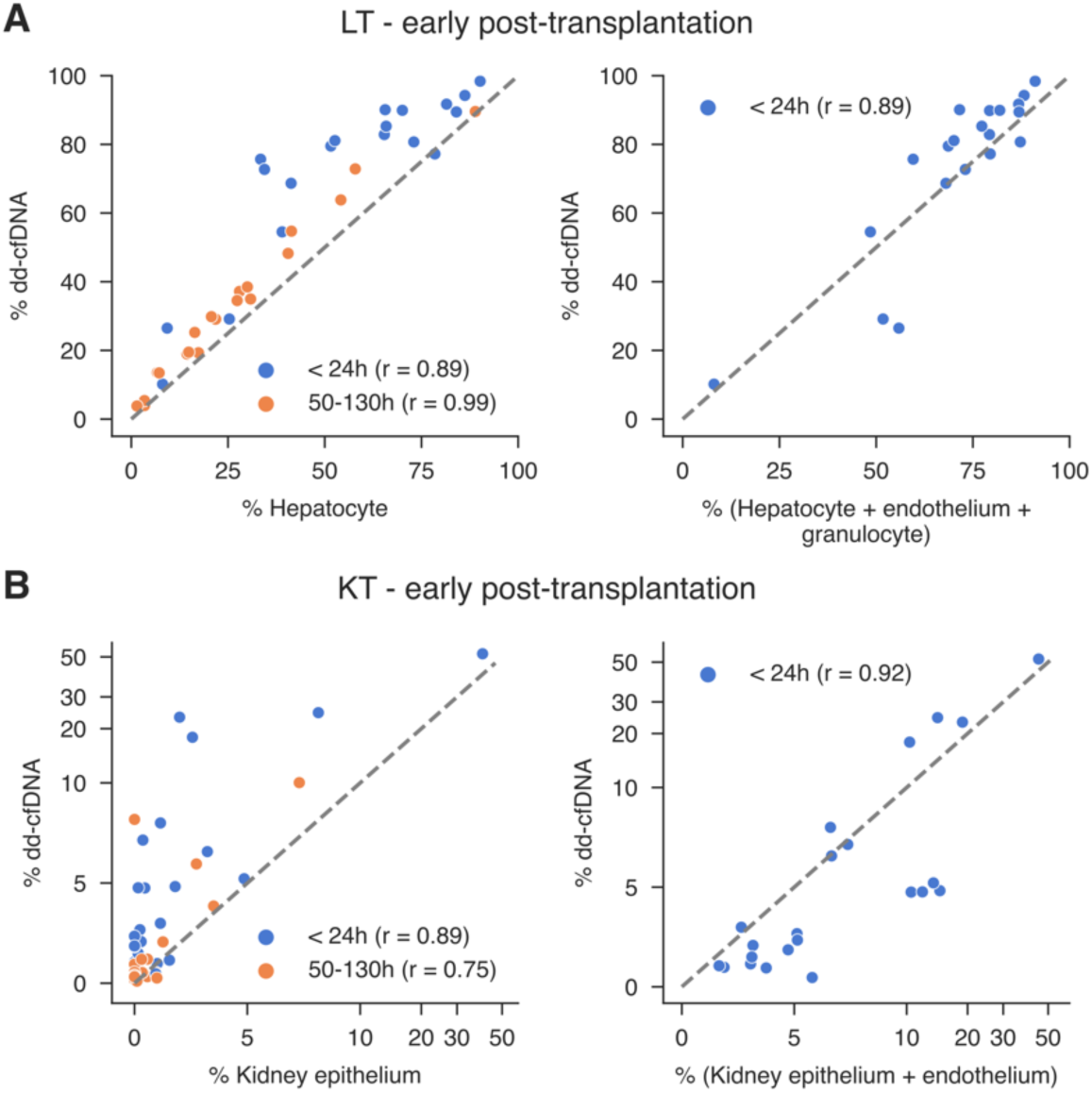
Correlation between plasma %dd-cfDNA and tissue-specific cfDNA fractions early post-transplantation. (**A**) Left: Correlation between plasma %dd-cfDNA and hepatocyte-derived cfDNA in LT recipients early post-transplant. Right: Correlation between %dd-cfDNA and the combined cfDNA fraction originating from hepatocytes, endothelium and granulocytes within 24 hours post-transplant. (**B**) Left: Correlation between %dd-cfDNA and kidney epithelium-derived cfDNA in KT recipients early post-transplant. Right: Correlation between %dd-cfDNA and the combined cfDNA fraction from kidney epithelium and endothelium within 24 hours. The x- and y-axes each use a hybrid scale: values below 10% are displayed on a linear scale, while values above 10% are displayed on a log10 scale. For all plots, each color represents a different collection time point. Pearson’s correlation coefficients are shown (all with P < 0.05). The grey line represents the identity line (y = x).

Notably, despite a median dd-cfDNA of 3.9% in the early phase, kidney epithelium accounted for only 0.98% (IQR: 0.18–2.0%) of total cfDNA. Several samples (n = 7) with dd-cfDNA in the range of 1.5– 7% exhibited kidney epithelium contributions of less than 0.5% (Figure 5B), with a median difference of all samples between %dd-cfDNA and kidney epithelium-derived cfDNA of 2.4% (IQR: 1.1–6.2%). This discrepancy was reduced in the later post-transplant period, where kidney epithelium-derived cfDNA more closely paralleled the overall %dd-cfDNA. In samples from the first 24 h post-transplantation, the addition of endothelium-derived cfDNA to kidney epithelium cfDNA yielded values that more closely correlated with the observed dd-cfDNA for samples with levels above 5% dd-cfDNA (Figure 5B). Unlike in LT recipients, granulocyte-derived cfDNA was not added to the endothelium fraction for KT recipients because their high median contribution of 38.5% (IQR: 28.2– 47.4%) to plasma cfDNA strongly outweighed the dd-cfDNA levels. Taken together, these results suggest that early post-transplantation dd-cfDNA in LT and KT patients is a composite reflecting variable contributions from multiple donor cell types over time.

Within the post-transplant cohort, approximately one-third (7/22) of KT recipients experienced delayed graft function (DGF), defined by the need for hemodialysis within the first week post-transplant (Table 1). Although DGF is linked to damage in kidney epithelium and endothelium (20), we observed no significant differences in the absolute quantities of cfDNA derived from these cell types or in dd-cfDNA levels between patients with and without DGF, regardless of collection time point. Similarly, no significant differences were observed when data were stratified by donor type or recipient sex. Also, no donor type- or recipient sex-specific differences were observed for hepatocyte- and endothelial-derived cfDNA in LT recipients.

## Discussion

This study demonstrates that our newly proposed targeted deep sequencing assay comprehensively evaluates cfDNA in transplant recipients by extending its analysis beyond traditional dd-cfDNA quantification. By incorporating TOO analysis, our multi-omics approach distinguishes the diverse cellular contributors to the circulating cfDNA pool in addition to quantifying dd-cfDNA, all while avoiding the need for resource-intensive whole-genome approaches. This advancement provides important new insights into cfDNA dynamics of potential relevance for transplant monitoring by identifying cfDNA release from numerous tissues, some of which are associated with common complications in solid organ transplantation.

We observed that stable liver and kidney allograft recipients exhibited a median of 3.5- to 4.5-fold higher total cfDNA levels than healthy individuals. This finding aligns with prior studies that reported similarly elevated cfDNA levels in long-term stable kidney (15) and heart transplant patients (21), potentially reflecting continuous cell turnover and increased injury to recipient tissue beyond the allograft. Based on these previous findings, we reasoned that a targeted assay designed to decipher the cfDNA TOO and quantify dd-cfDNA, as an established biomarker, might reveal the source of the increased recipient cfDNA and provide a more integrative view of the transplant patient’s health and cfDNA dynamics. A critical strength of our approach is its ability to reveal tissue-specific cfDNA release patterns that extend beyond the donor-derived signal and potentially indicate injury or increased cell turnover in other tissues. For instance, we observed that granulocyte-derived cfDNA was strongly increased in both stableLT and stableKT recipients compared to HC and was the dominant source of cfDNA in the stable phase. Similarly, cfDNA composition, particularly in granulocyte-derived cfDNA, also showed different dynamics between KT and LT recipients in the early post-transplant period. The lack of correlations between immune cell counts and their corresponding cfDNA levels suggests that these dynamics in cfDNA release are driven by altered cell turnover, immune responses or surgical trauma rather than corresponding changes in circulating cell count (18). One of the primary origins of granulocyte cfDNA is likely the formation of neutrophil or eosinophil extracellular traps (NET, EET), consistent with previous work showing increased neutrophil-derived cfDNA as a surrogate marker for NETosis (22,23). The observed increase in granulocyte-derived cfDNA in stable transplant recipients may indicate ongoing neutrophil activity, including increased NET formation. This finding aligns with the dual role of neutrophils in post-transplant processes. While crucial for tissue repair, excessive NET formation has been linked to graft dysfunction and rejection in both kidney and liver transplant recipients (24,25). Previous work has shown significantly increased levels of a marker for NETosis after transplantation in patients with previous end-stage renal disease (26). The elevated cfDNA levels from granulocytes may suggest a state of controlled neutrophil activation that, while not triggering overt rejection, may subtly influence graft microenvironments and potentially contribute to subclinical processes in stable transplant conditions.

Comparison of the dd-cfDNA fractions in the transplant recipients included here, with those reported in large-scale studies, showed findings fully consistent with a clinically stable post-transplant status. In stableKT recipients, stability is generally indicated by dd-cfDNA fractions below 0.5% (27). Our stableKT cohort had a median of 0.26%, a level consistent with patients who show no signs of subclinical injury (27). In contrast, stableLT recipients typically exhibit dd-cfDNA fractions below 10% (16), with our liver cohort having a median of 1.4%. These findings not only validate the accuracy of our measurements but also suggest that the increased recipient cfDNA from various cell types in our cohort is not driven by allograft injury. Interestingly, some of the tissue-specific cfDNA release patterns identified in our study are consistent with the increased risk for complications associated with immunosuppressive treatments. CNIs are known to be nephrotoxic, causing chronic injury to kidney tubular cells and exhibiting endothelial toxicity (17). The significant correlation between endothelial-derived cfDNA and cyclosporin A levels in our study may reflect CNI-induced vascular injury in these patients despite low dd-cfDNA levels. Elevated erythroid progenitor-derived cfDNA observed in KT recipients suggests differences in hematopoietic turnover between KT recipients and healthy individuals, in line with the common complication of post-transplant erythrocytosis in KT recipients due to persistently increased erythropoiesis (28). We also observed elevated pancreatic cfDNA in LT recipients potentially related to previously described CNI-induced endocrine toxicity (29,30) that may lead to new-onset diabetes mellitus after transplantation. Lastly, we demonstrated that hepatocyte cfDNA correlated with well-established biomarkers for liver injury in both the stable and early post-transplantation phases, supporting tissue-specific cfDNA as an injury marker. Differences in the strength of correlations between the different liver enzymes, particularly in the highly dynamic post-transplantation phase, may be related to the varying half-lives of these biomarkers.

For most stable LT recipients, the maintenance immunosuppression therapy consisted of either a CNI combined with mycophenolic acid/mycophenolate mofetil or a CNI combined with corticosteroids. In contrast, nearly all stable KT recipients received a triple-combination regimen comprising a CNI, mycophenolic acid/mycophenolate mofetil and corticosteroids, reflecting a more aggressive immunosuppressive approach. This difference in treatment may explain why stable LT recipients exhibit increased cfDNA release from certain cell types compared to stable KT recipients. More specifically, although the less intensive immunosuppression in stable LT recipients may be sufficient to maintain allograft function, the elevated cfDNA levels observed across various tissues could indicate a heightened subclinical inflammatory state inadequately reflected in CRP levels. Further investigation into a potential systemic inflammatory state in different organ transplant recipients and its relationship with immunosuppressive regimens is thus warranted.

In the early post-transplantation phase, we demonstrated for the first time that a substantial fraction of dd-cfDNA in LT and KT recipients does not solely originate from the primary transplanted tissue. Notably, immune cells and endothelium emerged as likely key contributors to the early post-transplant dd-cfDNA pool. Particularly in KT recipients, dd-cfDNA levels in the first 24 hours after transplantation were elevated with a subsequent decrease concordant with previous studies (14,19), whereas quantities of kidney epithelium-derived cfDNA were consistently low with no difference between the two time points. This observation that the primary transplanted tissue is not the major contributor to dd-cfDNA in KT recipients aligns with previous research (10), showing that following lung transplantation, small contributions from lung tissue are observed while most donor cfDNA was derived from immune cells. In KT recipients, the correlation with dd-cfDNA was improved when kidney epithelium cfDNA was combined with endothelial cfDNA. There are two potential explanations for this observation. On the one hand, much more limited epithelial damage might occur through reperfusion than previously assumed based on dd-cfDNA measurements, with endothelium being more strongly affected. Alternatively, cfDNA from kidney epithelium might predominantly be excreted through the urine, which could be investigated by TOO analysis of urinary cfDNA in future studies. Surprisingly, we did not observe a difference in graft tissue cfDNA between KT recipients with and without DGF despite its association with increased graft failure, acute allograft rejection and mortality one year post-transplantation (31). DGF is defined as the need for dialysis within the first week after transplantation. This subjective definition may partly explain the lack of difference observed, as it can vary considerably with center policies (32) and individual physician judgment. Alternatively, the overall low quantities of kidney epithelium-derived cfDNA in plasma early after transplantation, even in the presence of DGF, may reflect a predominant release of kidney epithelial cfDNA directly into the urine. Conversely to KT and lung transplantation (10), the majority of dd-cfDNA in LT recipients early after transplantation was derived from hepatocytes. Nevertheless, hepatocyte-derived cfDNA also did not account for all dd-cfDNA in LT. Although we cannot directly derive the TOO specifically for donor cfDNA fragments, the strong correlation between combined hepatocyte, endothelial and granulocyte cfDNA and dd-cfDNA proportions supports a model of multisource donor tissue cfDNA release also in LT recipients where endothelial damage and donor immune cell death may contribute significantly to early dd-cfDNA levels. Furthermore, by 50 to 130 hours after transplantation, the percentage of dd-cfDNA closely matched the proportion of hepatocyte cfDNA, in contrast to lung transplantation, where a significant contribution from non-lung tissues was reported to persist between 72 hours and 7 days (10). Finally, liver transplantation surgery, which typically involves more extensive skeletal muscle dissection and larger incisions than kidney transplantation, likely resulted in elevated levels of skeletal muscle cfDNA observed in LT but not KT recipients.

Additional consideration should be given to the possibility that our findings reflect a combination of altered cfDNA release from specific cell types and impaired cfDNA clearance. Clearance of circulating cfDNA primarily occurs through phagocytosis by Kupffer cells in the liver (33–35) and CNIs have been shown to impair Kupffer cell function (36), which could lead to reduced clearance of cfDNA in transplant recipients under immunosuppression. Similarly, the high number of cell types in LT recipients showing a significant decrease between the first 24 hours and 50 to 130 hours after transplantation, which was not observed in KT recipients, may partly be the result of decreased hepatic cfDNA clearance early after LT.

Our assay demonstrated robust analytical performance, as shown by in silico experiments, achieving high sensitivity for detecting graft tissue-specific cfDNA as low as 0.3% and dd-cfDNA as low as 0.1%. Dd-cfDNA measurements from our targeted deep sequencing assay strongly correlated with ddPCR measurements, confirming its accuracy relative to an established method (9,37,38). Our in silico experiments determined that a CV below 20% requires a certain level of sequencing depth. By applying our assay to over 150 plasma samples, we demonstrated that this depth of coverage can be reliably achieved from the input of a standard blood collection tube, underscoring its feasibility for clinical applications. Although current clinical thresholds indicating kidney allograft rejection remain well above 0.3%, a limitation of our assay is its lower detection limit of 0.3% for TOO analysis compared to dd-cfDNA, which resulted in no detection of kidney epithelium cfDNA in HC and stableKT recipients. Additionally, overlapping methylation signatures can lead to misclassification, as demonstrated by the detection of ovarian epithelium in some male participants, highlighting the potential benefit of further refinement of the cell type-specific markers in this assay. It is also important to note that while our assay validation relied on both in silico experiments and wet lab analyses, we did not assess TOO deconvolution specificity for all cell types. In addition, because some cell types, such as granulocytes, encompass multiple subpopulations with distinct functional roles, refining these markers to capture this heterogeneity may further enhance the diagnostic utility of cfDNA TOO analysis in transplant recipients and other clinical contexts. Future assay iterations could integrate additional markers beyond the initial 40 cell types, leveraging the expanding reference atlas data, to more accurately reflect the diversity of cell types contributing to cfDNA in patients with various pathologies.

In conclusion, this study shows that our targeted deep sequencing assay can expand cfDNA analysis by integrating TOO information, offering insights into transplant recipient health beyond dd-cfDNA. The assay showed robust analytical performance and identified distinct cfDNA release patterns associated with different transplanted organ types. This lays a promising foundation for future investigations into the potential of cfDNA analysis as an early indicator of complications in the absence of clinical presentation. Future research expanding this approach to the investigation of complications, including rejection and drug toxicity, could pave the way for more personalized risk stratification and management of organ transplant recipients. Finally, while this assay was applied in solid organ transplantation, TOO analysis is applicable in other clinical contexts to investigate the dynamics of tissue-specific cfDNA release and its potential as a diagnostic or prognostic biomarker.

## Methods

### Study cohort

Liver and kidney transplant recipients and healthy control individuals were recruited at the Inselspital, Bern University Hospital, Switzerland between 2020 and 2023. Exclusion criteria for transplant recipients were: being under 18 years of age, no blood sampling feasible at any of the intended time points and recipients of both kidney and liver or any other organ transplant. Exclusion criteria for healthy control participants were: being under 18 years of age, current or recent (within 3 months) pregnancy, a history of organ transplantation, current or recent (within 3 months) blood transfusion or administration of plasma-derived products, or having a current known malignancy.

Two cohorts of transplant recipients were included with kidney or liver allografts, one consisting of stable recipients, and the second cohort included patients with samples collected in the first week after transplantation (early post-transplantation cohort).

LT were deemed stable if they had not received any other organ transplanted than a liver, ASAT and ALAT were below 35 U/I (upper reference limit) and not more than two consecutive blood leukocyte measurements were above 10.5 x10^9^/L. KT recipients were deemed stable if they didn’t undergo dialysis treatment after 30 days post-transplantation, no indication biopsy was performed after 30 days post-transplantation, chronic kidney disease stage was lower than G3b, estimated glomerular filtration rate (eGFR) was > 30 mL/min./1.73m^2^ (allowing for a maximum of one measurement < 30) and a coefficient of variation < 1 for blood leukocyte measurements. Further, transplant recipients were excluded from the stable group if they underwent re-transplantation up to one year post-transplantation. All laboratory parameters were required to fulfill the criteria for measurements between 30 days post-transplantation and at least 1 month after sample collection.

Clinical data and laboratory parameters were extracted from the patients’ electronic health records. The time in hours after transplantation for the early-post transplantation cohort was calculated as the time since the final surgical suture timestamp, extracted from the patient’s electronic health record.

### Sample collection and cfDNA isolation

Venous blood samples were collected between the first day and up to 12 months after the transplantation from the transplant recipients. From the healthy control individuals, venous blood samples were collected at a single appointment. Up to 30 mL of whole blood from healthy controls and up to 15 mL from transplant recipients was collected in K3EDTA blood collection tubes (Sarstedt, Nümbrecht, Germany) and processed within 2 hours after collection. The blood samples were centrifuged at 2’000 g for 15 min. at room temperature (RT). The plasma was separated and re-centrifuged at 3’800 g for 10 min. at RT. The plasma supernatant and up to 1 mL of buffy coat were frozen following a pre-programmed freezing curve and subsequently stored at −80 °C in the Liquid Biobank Bern (www.biobankbern.ch) until DNA extraction.

For cfDNA isolation, the plasma was thawed at RT and isolated from 5 mL using the QIAamp Circulating Nucleic Acid Kit (Qiagen, Hilden, Germany). DNA concentration was measured using the Qubit dsDNA 1X HS Assay Kit (Thermo Fisher Scientific, Waltham, MA, USA).

### Primary human cell and genomic DNA isolation

Purified primary human cells from the liver, kidney and immune system were used to validate the targeted methylation panel. Cryopreserved purified hepatocytes in suspension from a single female donor were ordered from Biopredict International (Saint-Grégoire, France). Purified human renal cortical epithelial cells (HRCEpC) from a single male donor suspended in RNAlater (Thermo Fisher Scientific, Waltham, MA, USA) were obtained from PromoCell (Heidelberg, Germany). B cells were isolated from whole blood of two healthy control individuals using Dynabeads™ CD19 Pan B (Thermo Fisher Scientific, Waltham, MA, USA). T cells were isolated from whole blood of a healthy control individual using Dynabeads™ HLA Class I (Thermo Fisher Scientific, Waltham, MA, USA). The HLA Class I beads specifically bind to the CD8 membrane antigen on human cells and hence isolate CD8^+^T lymphocytes. The differential CBC of whole blood from healthy control individuals was determined using the XN-1000 Hematology Analyzer (Sysmex Suisse AG, Horgen, Switzerland) before genomic DNA (gDNA) extraction. The buffy coat was removed from the ETDA blood sample from healthy control individuals after the first centrifugation at 2’000 g. The buffy coat was washed twice by resuspension in 10 mL PBS (no Mg^2+^ or Ca^2+^) and subsequently pelleted at 300 g for 15 min., followed by a final resuspension in 1.5 mL of PBS. The gDNA was extracted from 200 µl whole blood, the buffy coat cell suspensions and suspensions of cells bound to the Dynabeads™ using the QIAamp DNA Blood Mini Kit (Qiagen, Hilden, Germany). DNA concentration was measured using the Qubit dsDNA 1X HS Assay Kit (Thermo Fisher Scientific, Waltham, MA, USA).

The gDNA was sheared in 20 μl 10 mM Tris-Cl (pH 8.5) to 170 bp using the COVARIS LE220+ ultrasonicator (Covaris Inc., Woburn, MA, USA). The sheared gDNA from the pure cell types was size selected to resemble the average profile of cfDNA. For that, Clarefy DNA purification beads (Claret Bioscience, Santa Cruz, CA, USA) were added at 0.7x ratio, incubated for 10 min. and placed on a magnetic rack. The supernatant was transferred to a new tube, beads were added at a 1.2x ratio, incubated for 10 min. and placed on a magnetic rack. The supernatant was discarded, the beads were washed twice with 80% ethanol and the DNA was eluted in 32 µl nuclease-free water. DNA concentration was measured using the Qubit dsDNA 1X HS Assay Kit (Thermo Fisher Scientific, Waltham, MA, USA).

### Panel design

A hybridization capture panel was designed to target cell type-specific hypomethylated regions for TOO analysis and SNPs for dd-cfDNA quantification. The panel design for TOO analysis was based on cell-type-specific markers for 40 different cell types published by Loyfer et al. (11), including data from 39 cell types and megakaryocyte-specific data (39). The markers were selected as blocks of CpGs with similar methylation levels across three or more CpGs in the human genome that are differentially methylated between the target and the background cell types.

The panel was developed using a stepwise design approach: For the methylation marker selection, cell type-specific markers were ranked as previously described (11) based on the difference in the average methylation between the target cell type and other cell types. Markers were filtered to exclude regions with complete overlap with Alu repeat annotations (40,41) to minimize off-target binding of the capture probes. SNPs were selected for dd-cfDNA fraction determination based on the following criteria: 1. minor allele frequency (MAF) between 0.45 and 0.55 (42), 2. absence of G and C alleles in the databases (42), 3. non-overlapping with short interspersed nuclear elements (SINEs) annotations (43) and 4. distributed across all 22 autosomes.

Capture probes were designed by Twist Bioscience (South San Francisco, CA, USA) against hypermethylated (100% methylated CpGs) and hypomethylated (100% CpGs) fragments from the target regions focusing on achieving high coverage of the marker CpG sites. We further refined the design by evaluating the coverage of the markers by probes using varying probe tiling densities (1x, 1.5x and 2x). This process aimed to optimize the trade-off between maximizing CpG site coverage and minimizing redundant capture of non-essential regions. A tiling density of 1.5x was chosen. To evaluate the design, the regions were manually inspected and markers where CpG sites were inadequately covered by probes, assuming a mean library insert size of 167 bp, were omitted from the panel. All SNPs were fully covered by probes.

The NGS Methylation Detection System (Twist Bioscience, South San Francisco, CA, USA) platform was used and 120 bp biotinylated probes were synthesized by Twist Bioscience. Probes with increased off-target rates were excluded from the final panel design.

The final panel design targeted 975 cell type-specific markers (Supplementary Data) with a median marker length of 283 bp (IQR: 163–445 bp) and a median of 7 CpGs (IQR: 6–10 CpGs) per marker. Regarding the SNPs, the first version of the panel targeted 117 SNPs, with 115 on all 22 autosomes and two on the X chromosome. A second version of the panel was designed that included 302 SNPs (Supplementary Data), with 300 on all 22 autosomes and two with one on each sex chromosome. While some samples were analyzed with the first version of the panel and the rest with the second version, equivalent performance was shown (Supplementary Figure 7). The SNPs on the sex chromosomes were not used for %dd-cfDNA quantification but for quality control purposes only. The final panel design covered a total target region of approximately 497 kbp.

### ddPCR dd-cfDNA and total cfDNA quantification

The fractional abundance and absolute GE/mL plasma of dd-cfDNA were determined as previously described (38) using ddPCR and assays targeting mismatches between the donor and recipient in the *HLA-DRB1* gene. The total amount of cfDNA was determined by measuring the copy number of the gene *RPP30* using a previously published assay (44) following the same ddPCR protocol as for the *HLA-DRB1* assays.

### Targeted methylation sequencing

For sequencing, up to 150 ng of sheared gDNA or up to 70 ng of cfDNA was converted using the NEBNext Enzymatic Methyl-seq Conversion Module (New England Biolabs, Ipswich, MA, USA) by oxidizing 5-mC to 5-hmC and subsequently deaminating unmethylated cytosines. The clean-up steps were performed using Clarefy DNA purification beads (Claret Bioscience, Santa Cruz, CA, USA). The enzymatic conversion protocol was adapted to maximize recovery by adding 100 µl of purification beads for the first and 170 µl for the second clean-up. The DNA was eluted in 16 µl nuclease-free water after the first clean-up. The converted DNA after the second clean-up was eluted in 18 μl and the whole eluate was immediately used as input for library preparation with the SRSLY NanoPlus kit (Claret Bioscience, Santa Cruz, CA, USA). Unique molecular identifiers (UMI) and unique dual indices (UDI) were used, and 8–10 index PCR cycles were performed depending on the original input as instructed in the manual. The DNA library concentration was quantified using the Qubit dsDNA 1X HS Assay Kit (Thermo Fisher Scientific, Waltham, MA, USA), while the quality and fragment size distribution were analyzed with the Agilent High Sensitivity DNA Kit (Agilent Technologies Inc., Santa Clara, CA, USA). The libraries were stored at −20 °C until target enrichment. Up to 14 libraries were multiplexed and target enrichment was performed with up to 2 µg total input per reaction using the Twist NGS Methylation Detection System (Twist Bioscience, San Francisco, CA, USA). The probes were hybridized to the targets for two hours and 15 post-capture PCR cycles were performed. Final concentration, quality and fragment size were determined as described above. Up to five enriched library pools were combined, spiked with PhiX v3 library (Illumina, San Diego, CA, USA) at 10% and sequenced with 150 bp paired-end reads on a NextSeq 550 or NovaSeq 6000 (Illumina, San Diego, CA, USA).

### Bioinformatic analysis

Demultiplexing and UMI sequence extraction were performed using SRSLYumi v0.3 (45) and the bcl2fastq v2.20.0 software. Adapter and quality trimming was performed with Trim Galore v0.6.10 (46) and the reads were mapped to the human genome hg19 using bwa-meth v0.2.6 (47). The mapped reads were grouped based on their genomic coordinate and UMI with UMI-tools v1.1.4 (48) and consensus reads were created from groups with one or more reads using fgbio v2.1.0 (49). The consensus reads were remapped, filtered (mapping quality ≥50), stripped from non-CpG nucleotides and converted to PAT and BETA files using wgbstools v0.2.1 (50). The TOO deconvolution was performed using a fragment-level non-negative least squares-based algorithm as previously described (11) using reads with ≥ 4 CpGs.

The cytosine conversion efficiency was determined based on the average methylation rate of non-CpG-context cytosines. The target region coverage was determined using Picard tool’s CollectHsMetrics v3.0.0 (51).

To determine the fraction of dd-cfDNA, the number of filtered consensus reads containing each allele per SNP was calculated using SAMtools mpileup (52). The %dd-cfDNA for each SNP was computed as the count of the alternate allele divided by the total allele count. For samples with dd-cfDNA fractions exceeding 20%, the recipient’s genotype was employed to identify and exclude heterozygous SNPs in the recipient. Subsequently, the recipient’s genotype was utilized to identify the donor allele for each SNP where the recipient was homozygous and a second allele was detected. The recipient genotype was determined by sequencing gDNA from the recipient’s buffy coat. For fractions ≤ 20%, SNPs with %dd-cfDNA between 30% and 70% were assumed to be recipient heterozygous and excluded. Outliers, defined as SNPs with %dd-cfDNA > 2 standard deviations from the mean %dd-cfDNA, were removed. For samples with a mean %dd-cfDNA > 10%, donor genotypes were incorporated to adjust estimates by identifying donor homozygous and heterozygous SNPs. To identify donor genotypes, a gaussian mixture model assuming the three components was applied, implemented in scikit-learn v1.3.0, enabling the classification of SNPs as donor homozygous for the reference allele, homozygous for the alternate allele or heterozygous. The mean of SNPs where both donor and recipient were homozygous for the same allele was subtracted from the adjusted mean to account for background.

Absolute GE/mL for cfDNA TOO and dd-cfDNA were determined by multiplying the cfDNA concentration (ng/mL) by a factor of 303, derived from the haploid human genome weight in pg.

### In silico mixtures

For TOO analysis based on in silico mixtures, hepatocyte gDNA reads were combined with a background of WBC gDNA reads from three healthy control individuals. In contrast, kidney gDNA reads were mixed into a mixture consisting of 97% WBC and 3% hepatocyte gDNA reads (for a plasma-like background). A shuffled subset of both hepatocyte and kidney gDNA reads were mixed into the background at various fractions ranging from 90% to 0%. All mixtures were generated with a target region coverage ranging from 30x to 1000x. For each mixture and coverage combination, 20 replicates were generated. Read mixing was performed using wgbstools v0.2.1 (50). For the SNP-based dd-cfDNA quantification in silico mixtures, plasma cfDNA reads from one healthy control individual sample were mixed into a background of reads from a second healthy control individual sample at various fractions between 90% and 0%. The target region coverage ranged from 200x to 1000x. For each mixture and coverage combination, 20 replicates were generated. Read mixing was performed using SAMtools (52).

### Statistics

All statistical analyses and graphs were performed and created using Python v3.10. Global statistical testing per cell type between three groups was conducted using the non-parametric Kruskal-Wallis test (SciPy v1.10) with P values adjusted across cell types using the Benjamini-Hochberg procedure (statsmodels v0.13.5), with the false discovery rate (FDR) threshold set at 0.05 to control for the type I error. For significant global tests, the Dunn’s post-hoc test was employed to identify pairwise differences. Dunn’s post-hoc P values were adjusted for multiple testing across groups using the Bonferroni method (statsmodels v0.13.5). To assess differences between two groups, the Wilcoxon signed-rank test (statsmodels v0.13.5) was utilized for paired samples, while the Mann–Whitney U test (statsmodels v0.13.5) was applied for unpaired samples. Multiple hypothesis testing, without preceding global testing, was corrected using the Benjamini-Hochberg procedure (statsmodels v0.13.5). P values were deemed significant when less than 0.05. For correlation analysis, the Pearson’s correlation coefficient was calculated.

## Data Availability

Raw adapter trimmed FASTQ files generated in this study have been deposited at the European Genome-phenome Archive (EGA), which is hosted by the EBI and the CRG, under accession number EGAS50000000987. Due to Swiss law, access to the data is restricted. Researchers wishing to obtain the data must submit an application to the responsible Data Access Committee.

## Declarations

### Ethics approval and consent to participate

The study was approved by the ethics committee of the Canton of Bern, Switzerland (2020–00953, 2019–00730), with procedures performed in accordance with the Declaration of Helsinki. Blood samples were obtained from participants who have provided written informed consent.

### Availability of data and materials

Raw adapter trimmed FASTQ files generated in this study have been deposited at the European Genome-phenome Archive (EGA), which is hosted by the EBI and the CRG, under accession number EGAS50000000987. Due to Swiss law, access to the data is restricted. Researchers wishing to obtain the data must submit an application to the responsible Data Access Committee. For the preprocessing, an analysis pipeline was implemented in Snakemake v7.22.0 (53) and is available at https://github.com/NichKu/TransMethylCFDx. The scripts used for dd-cfDNA calculation and in silico mixing were also made available there.

### Author’s contributions

N.K. and U.A. designed the study. N.K. and F.S. recruited healthy control participants. D.S., V.B. and A.B. recruited patients with the help of F.S., while F.S. collected the clinical data. N.K. performed the research, acquired the data and performed the data analysis. N.K. wrote the manuscript with U.A. C.K.Y.N. and C.R.L. provided scientific advice and helped with the interpretation of the data. All authors reviewed and approved the final version of the manuscript.

## Acknowledgments

The authors would like to thank all the patients and their families, as well as the healthy control volunteers, for their participation. We further thank all the University Institute of Clinical Chemistry and Clinical Genomics Lab technicians and staff of the Inselspital for their laboratory support, namely, Daniel Schärer, Gisela Andrey and Nicole Klaus. We also thank all nurses and medical staff involved in the sample collection and appreciate the support of Netanel Loyfer, who was the lead author of the publication on the methylation atlas. The study overview figure (Figure 1) in this manuscript was created with BioRender.com.

