## Supplementary Figures for "Integrated targeted deep sequencing reveals unique tissue-of-origin and donor-derived cell-free DNA signatures in organ transplant recipients"

Nicholas Kueng[1,2], Fanny Sandberg[1,2], Daniel Sidler[3], Vanessa Banz[4], Annalisa Berzigotti[5], Charlotte K. Y. Ng[6,7], Carlo R. Largiadèr[1], Ursula Amstutz[1]*

### Affiliations
[1] Department of Clinical Chemistry, Inselspital, Bern University Hospital, University of Bern, Bern, Switzerland
[2] Graduate School for Cellular and Biomedical Sciences, University of Bern, Bern, Switzerland
[3] Department of Nephrology and Hypertension, Inselspital, Bern University Hospital and University of Bern, Bern, Switzerland
[4] Department of Visceral Surgery and Medicine, Inselspital, Bern University Hospital and University of Bern, Bern, Switzerland
[5] Department of Hepatology, Inselspital, Bern University Hospital and University of Bern, Bern, Switzerland
[6] IRCCS Humanitas Research Hospital, Milan, Italy
[7] Humanitas University, Milan, Italy

*Corresponding author

### Table of Content

Supplementary Figure 1 - Tissue-of-origin deconvolution results of in silico mixtures at 30x sequencing depth.

Supplementary Figure 2 - Linearity of tissue-of-origin deconvolution.

Supplementary Figure 3 - Linearity of dd-cfDNA quantification.

Supplementary Figure 4 - Coefficient of variation for in silico mixtures for different tissue proportions and target region depth of coverage.

Supplementary Figure 5 - Coefficient of variation for in silico mixtures for different donor-derived cfDNA proportions and target region depth of coverage.

Supplementary Figure 6 - Agreement of Qubit- vs ddPCR-based total cfDNA quantification.

Supplementary Figure 7 - Comparability of dd-cfDNA quantification with two SNP panel versions.

Supplementary Figure 8 - Absolute cfDNA per cell type for stable transplant recipients versus healthy controls excluding granulocyte cfDNA.

Supplementary Figure 9 - %dd-cfDNA in stable transplant recipients.

Supplementary Figure 10 - Absolute cfDNA of non-hematological origin in stable transplant recipients versus healthy controls.

Supplementary Figure 11 - Absolute cfDNA tissue-of-origin from all 40 cell types for stable transplant recipients versus healthy controls.

Supplementary Figure 12 - Correlation between absolute endothelium cfDNA versus matched plasma cyclosporin A levels.

Supplementary Figure 13 - Correlation between absolute hepatocyte cfDNA versus liver enzyme measurements in the stable transplant cohorts.

Supplementary Figure 14 - Total cfDNA early post-transplantation.

Supplementary Figure 15 – Absolute cfDNA tissue-of-origin from all 40 cell types for kidney transplant recipients at two time points early post-transplantation.

Supplementary Figure 16 – Absolute cfDNA tissue-of-origin from all 40 cell types for liver transplant recipients at two time points early post-transplantation.

Supplementary Figure 17 - Correlation between absolute hepatocyte cfDNA and liver enzyme measurements in liver transplant recipients from the early post-transplantation cohort.

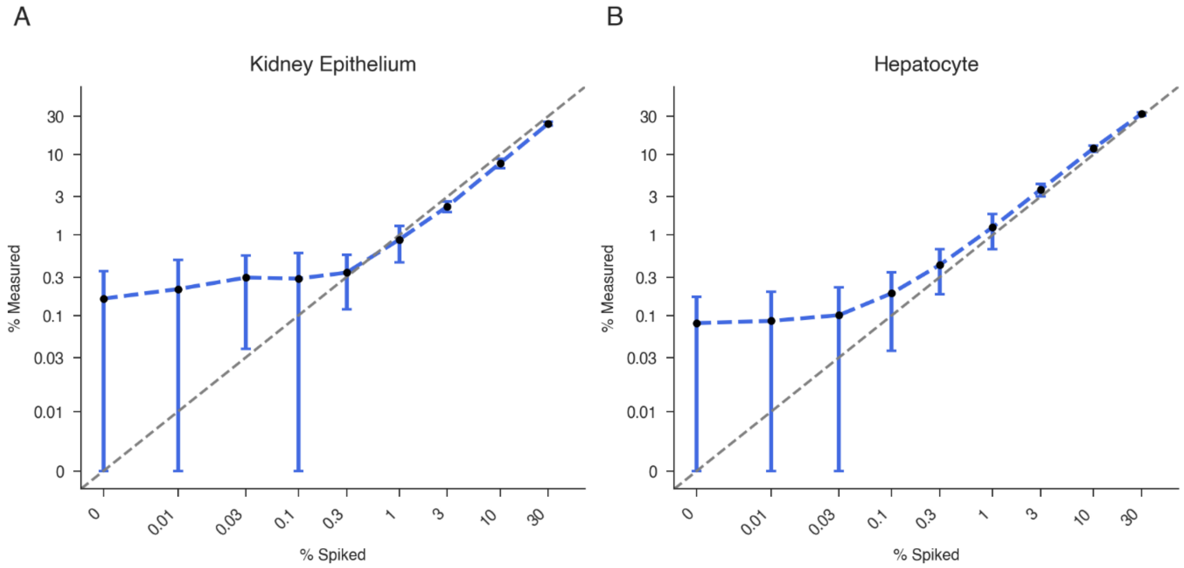


**Supplementary Figure 1: Tissue-of-origin deconvolution results of in silico mixtures at 30x sequencing depth.**
Reads from sequenced genomic DNA from kidney epithelium (**A**) or hepatocyte (**B**) samples were computationally mixed into a background of leukocyte genomic DNA reads from three healthy control individuals. The background for the kidney epithelium (**A**) mixtures consisted of 97% leukocyte and 3% hepatocyte reads. Mixtures were performed at a sequencing depth of 30x. Black markers show the median determined contribution for 20 replicates with the error bars displaying one standard deviation. The grey line (where displayed) represents the identity line (y=x).

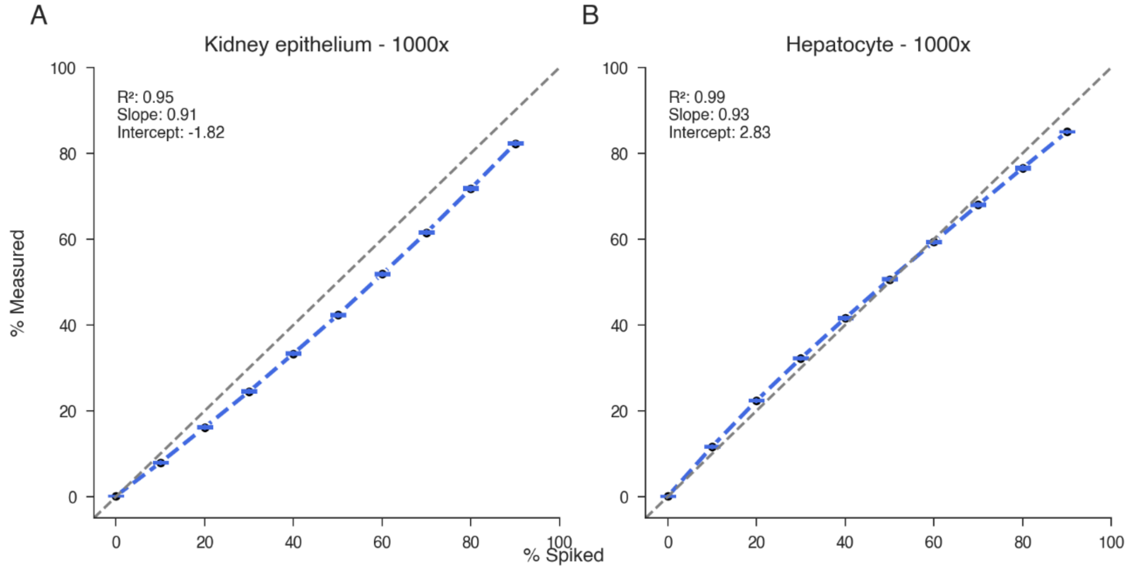


**Supplementary Figure 2: Linearity of tissue-of-origin deconvolution.**
Reads from sequenced genomic DNA from kidney epithelium (**A**) or hepatocytes (**B**) samples were computationally mixed into a background of leukocyte genomic DNA reads from three healthy control individuals. The background for the kidney epithelium (**A**) mixtures consisted of 97% leukocyte and 3% hepatocyte reads. Mixtures were performed at a sequencing depth of 1’000x. Black markers show the median determined contribution for 20 replicates with the error bars displaying one standard deviation. The grey line represents the identity line (y=x).

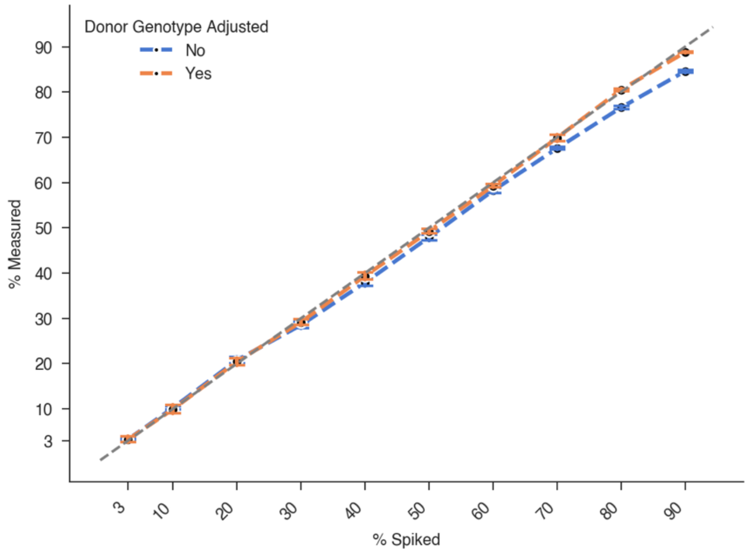


**Supplementary Figure 3: Linearity of dd-cfDNA quantification.**
Plasma cfDNA reads from two healthy controls were mixed at various fractions. Mixture was performed at a sequencing depth of 1’000x. Black markers show the median determined contribution for 20 replicates with the error bars displaying one standard deviation. The grey line represents the identity line (y=x).

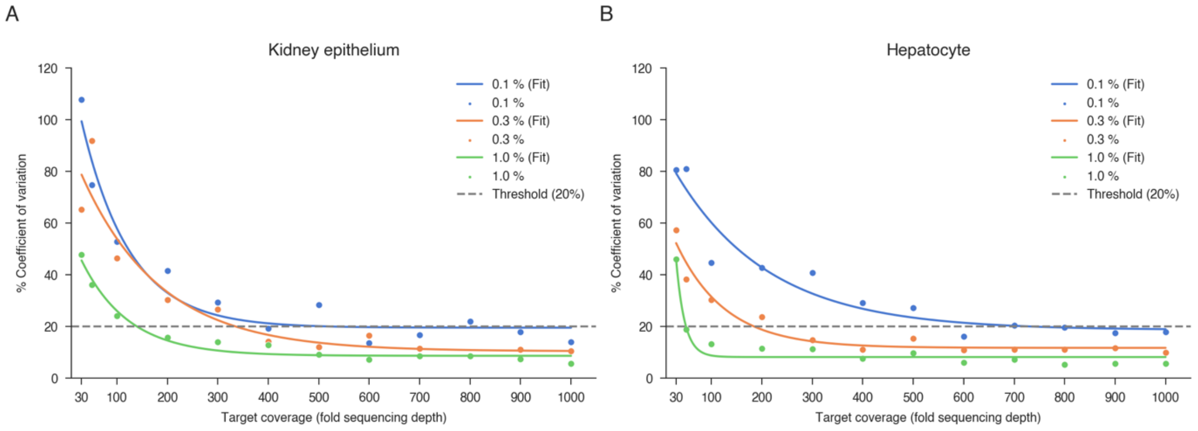


**Supplementary Figure 4: Coefficient of variation for in silico mixtures for different tissue proportions and target region depth of coverage.**
Reads from sequenced genomic DNA from kidney epithelium (**A**) or hepatocytes (**B**) samples were computationally mixed into a background of leukocyte genomic DNA reads from three individuals at 0.1, 0.3 and 1.0% fractions. The background for the kidney epithelium (**A**) mixtures consisted of 97% leukocyte and 3% hepatocyte reads. The mixtures were produced for different target region depth of coverage (x-axis). The y-axis indicates the coefficient of variation (CV), calculated across 20 replicates. The dashed grey line represents the threshold for a CV of 20%.

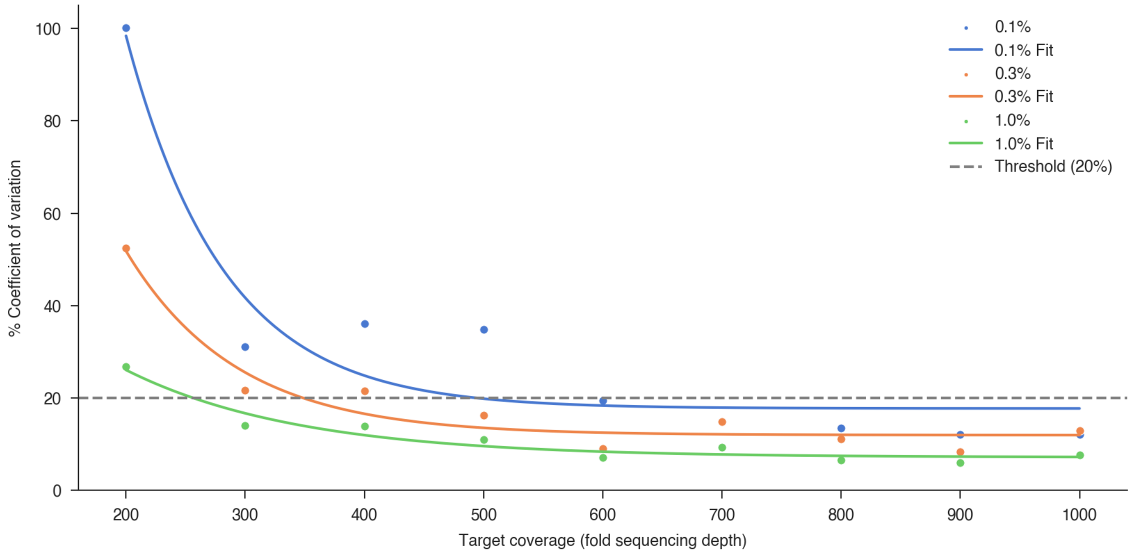


**Supplementary Figure 5: Coefficient of variation for in silico mixtures for different donor-derived cfDNA proportions and target region depth of coverage.**
Plasma cfDNA reads from two healthy controls were mixed at 0.1, 0.3 and 1.0% fractions. The mixtures were produced for different target region depth of coverage (x-axis). The y-axis shows the coefficient of variation (CV), calculated across 20 replicates. The grey line represents the threshold for a CV of 20%.

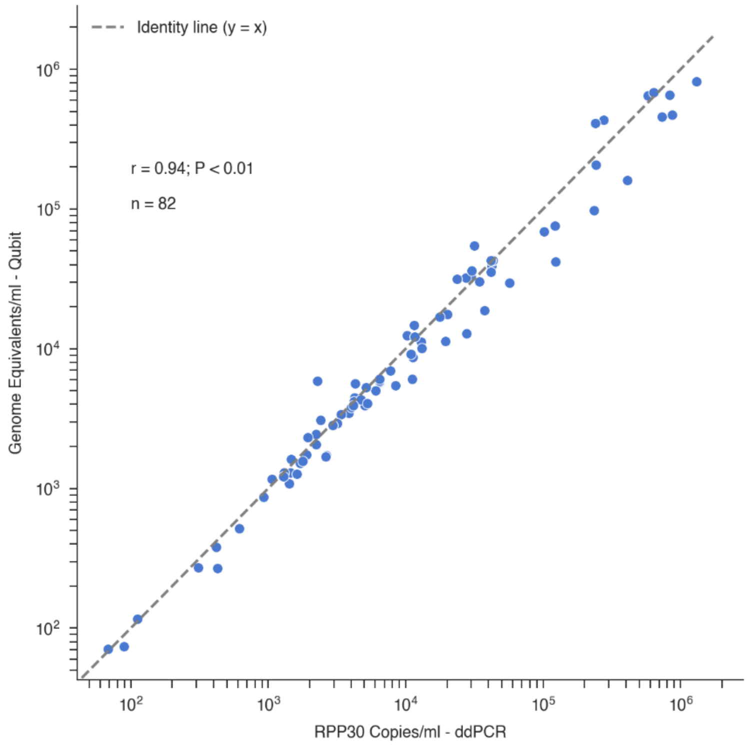


**Supplementary Figure 6: Agreement of Qubit- vs ddPCR-based total cfDNA quantification.**
Scatterplot of plasma cfDNA concentrations (n = 82 samples) quantified with Qubit and ddPCR. *RPP30* gene copies were measured with ddPCR. Pearson's r correlation coefficient is shown. The grey line represents the identity line (y=x).

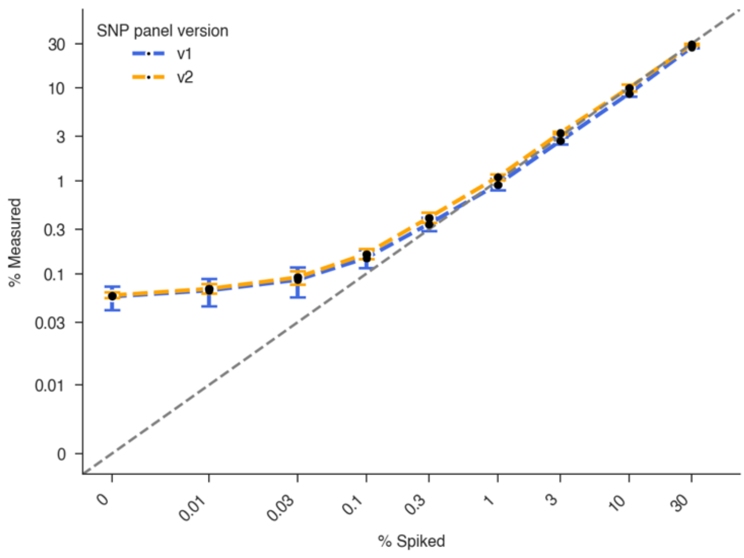


**Supplementary Figure 7: Comparability of dd-cfDNA quantification with two SNP panel versions.**
Plasma cfDNA reads from two healthy controls were mixed at various fractions. Mixtures were performed at a sequencing depth of 1’000x. Dd-cfDNA fractions were calculated based on the SNPs included either in panel version v2 or with the selection of SNPs from v1, a subset of v2. Black markers show the median determined dd-cfDNA fraction for 20 replicates with the error bars displaying one standard deviation. The grey line represents the identity line (y=x).

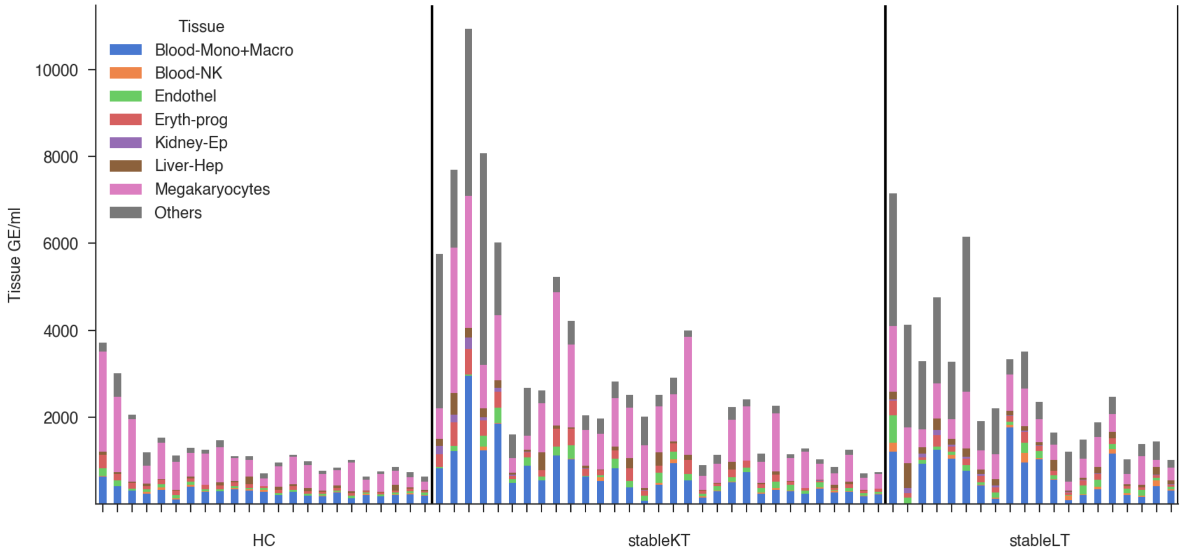


**Supplementary Figure 8: Absolute cfDNA per cell type for stable transplant recipients versus healthy controls excluding granulocyte cfDNA.**
Absolute tissue-of-origin GE/mL per individual sample, with bars grouped by cohort and in descending order of total GE/mL. Each colored segment represents a distinct cell type (without granulocytes) or cell type group. StableLT: stable liver transplant recipients (n = 20), stableKT: stable kidney transplant recipients (n = 31), HC: healthy control group (n = 23).

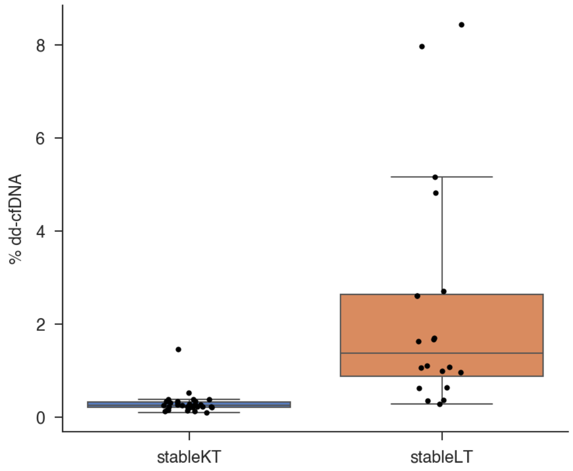


**Supplementary Figure 9: %dd-cfDNA in stable transplant recipients.**
Boxplot illustrating the %dd-cfDNA levels in stable transplant recipients. StableLT: stable liver transplant recipients (n = 20), stableKT: stable kidney transplant recipients (n = 31). The box spans the 25th to 75th percentiles with a line at the median. Whiskers extend to 1.5× the interquartile range.

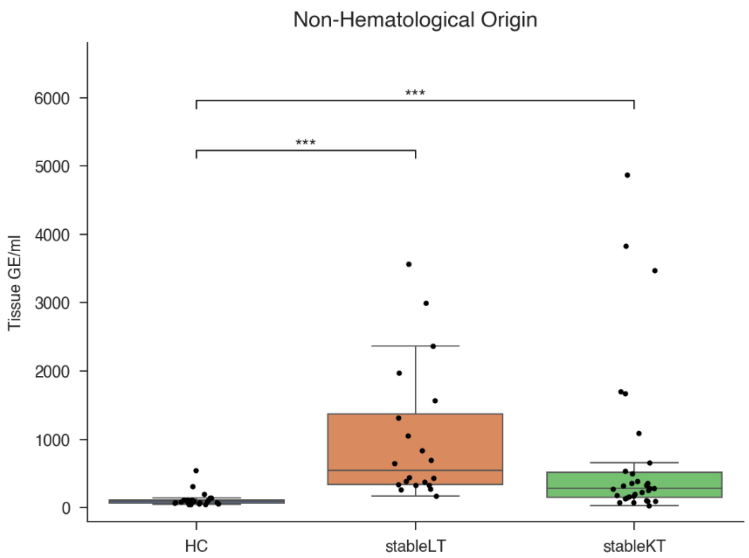


**Supplementary Figure 10: Absolute cfDNA of non-hematological origin in stable transplant recipients versus healthy controls.**
Boxplot illustrating the absolute cfDNA levels from non-hematological sources across different groups. StableLT: stable liver transplant recipients (n = 20), stableKT: stable kidney transplant recipients (n = 31), HC: healthy control group (n = 23). Kruskal-Wallis test was used for global testing and Dunn's post-hoc test for multiple pairwise comparisons between the groups. Dunn's test P values were adjusted using the Bonferroni correction to control for multiple comparisons. *** $P < 0.001$, no comparison annotation means $P > 0.05$. The box spans the 25th to 75th percentiles with a line at the median. Whiskers extend to 1.5× the interquartile range.

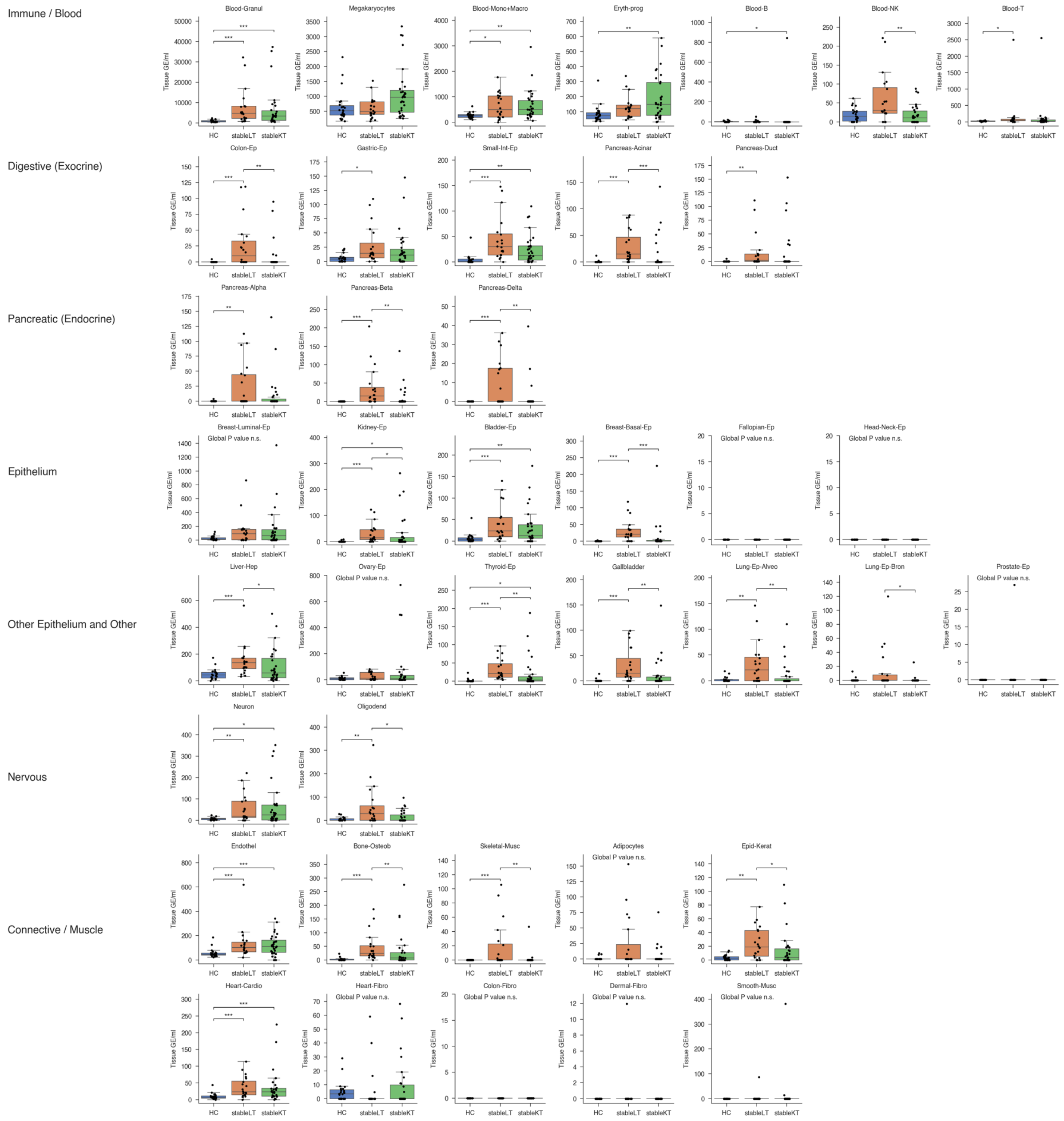


**Supplementary Figure 11: Absolute cfDNA tissue-of-origin from all 40 cell types for stable transplant recipients versus healthy controls.**

Boxplots of absolute cfDNA from all cell types by cohort group. StableLT: stable liver transplant recipients (n = 20), stableKT: stable kidney transplant recipients (n = 31), HC: healthy control group (n = 23). For each cell type, the Kruskal-Wallis test was performed for global comparisons, with the Benjamini-Hochberg procedure applied to control the false discovery rate (FDR) across cell types. For significant results, Dunn's post-hoc test was used for pairwise comparisons between groups, and its P values were adjusted using the Bonferroni correction to account for multiple comparisons between groups for a given cell type. * P < 0.05, ** P < 0.01, *** P < 0.001, **** P < 0.0001, no comparison annotation means P > 0.05. n.s. means not statistically significant. The box spans the 25th to 75th percentiles with a line at the median. Whiskers extend to 1.5x the interquartile range.

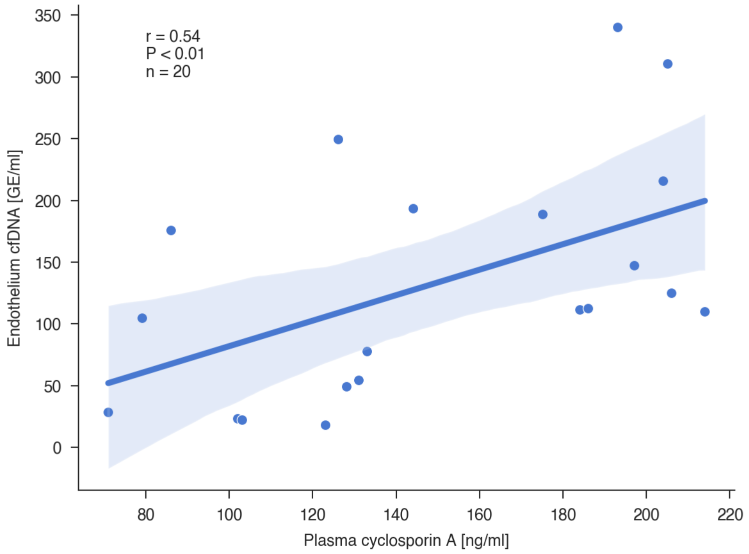


**Supplementary Figure 12: Correlation between absolute endothelium cfDNA versus matched plasma cyclosporin A levels.**
The blue line represents the linear regression with its corresponding 95%-confidence interval. Shown is the Pearson's r correlation coefficient.

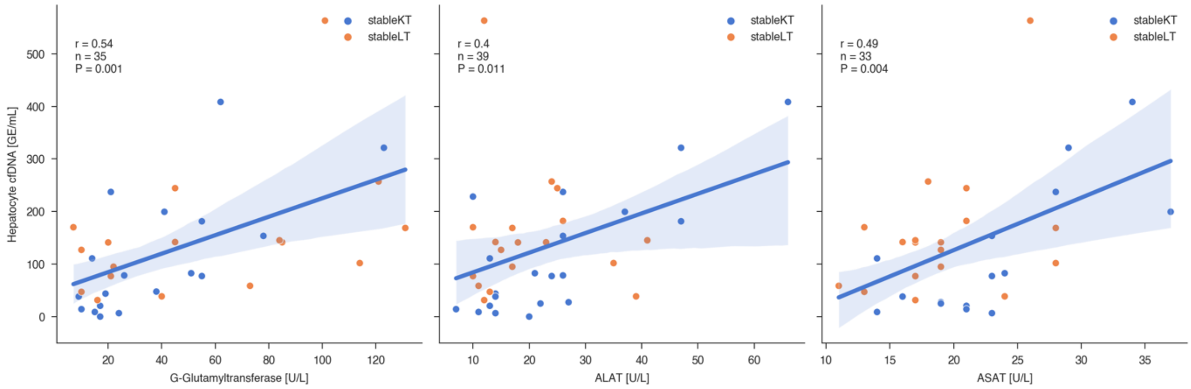


**Supplementary Figure 13: Correlation between absolute hepatocyte cfDNA versus liver enzyme measurements in the stable transplant cohorts.**
The blue lines represent the linear regression with its corresponding 95%-confidence interval. Shown is the Pearson's r correlation coefficient. stableLT: stable liver transplant recipients, stableKT: stable kidney transplant recipients.

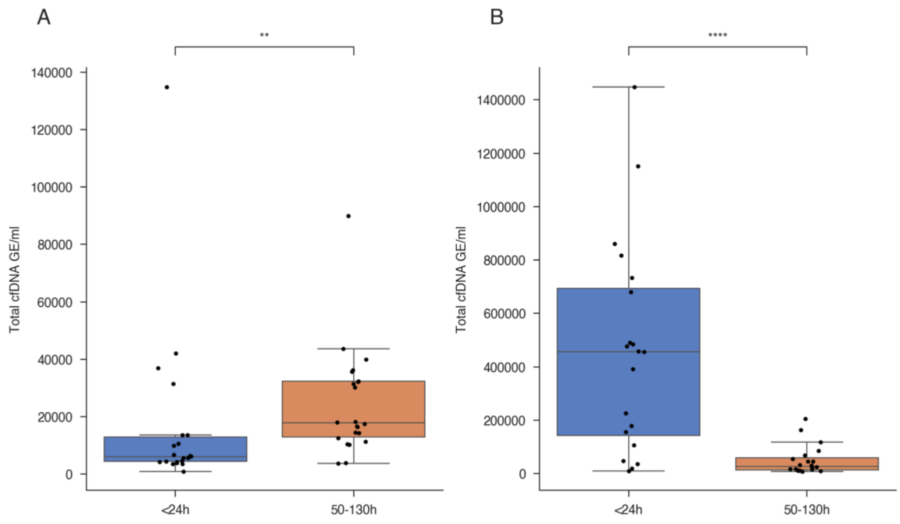


**Supplementary Figure 14: Total cfDNA early post-transplantation.**
Boxplot of total cfDNA for (**A**) kidney transplant (n = 22) and (**B**) liver transplant recipients (n = 20) early post-transplantation. Time on the x-axis indicates the time since the transplantation in which the samples were collected. The Wilcoxon signed-rank test was used. ** $P < 0.01$, *** $P < 0.001$. The box spans the 25th to 75th percentiles with a line at the median. Whiskers extend to 1.5× the interquartile range.

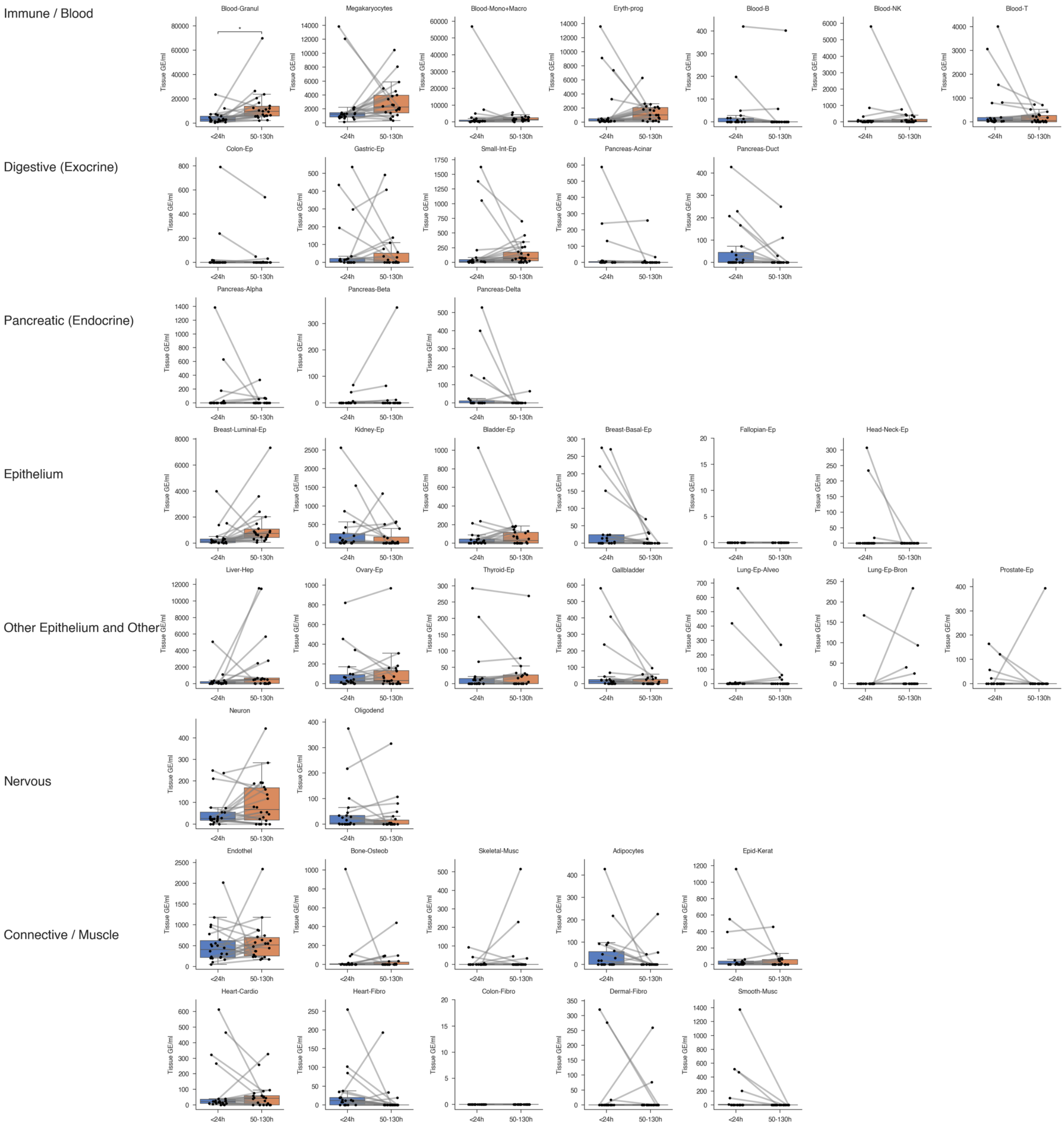


**Supplementary Figure 15: Absolute cfDNA tissue-of-origin from all 40 cell types for kidney transplant recipients at two time points early post-transplantation.**

Boxplot of absolute cfDNA from all cell types by time point. The Wilcoxon signed-rank test was used, and P values were adjusted for multiple comparisons across cell types using the Benjamini-Hochberg procedure to control the false discovery rate (FDR). * $P < 0.05$, no comparison annotation means $P > 0.05$. The box spans the 25th to 75th percentiles with a line at the median. Whiskers extend to 1.5× the interquartile range.

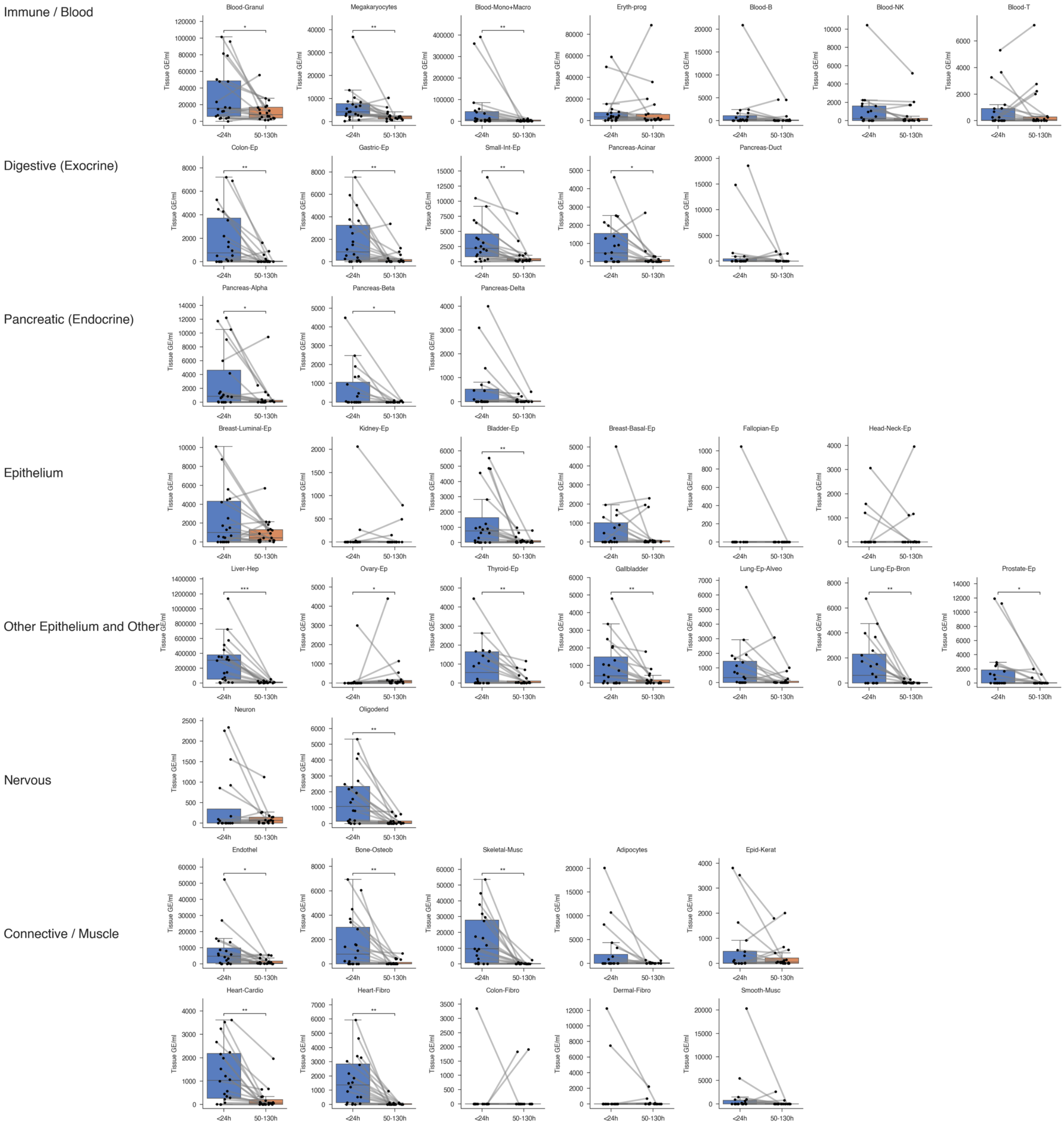


**Supplementary Figure 16: Absolute cfDNA tissue-of-origin from all 40 cell types for liver transplant recipients at two time points early post-transplantation.**

Boxplot of absolute cfDNA from all cell types by time point. The Wilcoxon signed-rank test was used, and P values were adjusted for multiple comparisons across cell types using the Benjamini-Hochberg procedure to control the false discovery rate (FDR). * P < 0.05, ** P < 0.01, *** P < 0.001, **** P < 0.0001, no comparison annotation means P > 0.05. The box spans the 25th to 75th percentiles with a line at the median. Whiskers extend to 1.5× the interquartile range.

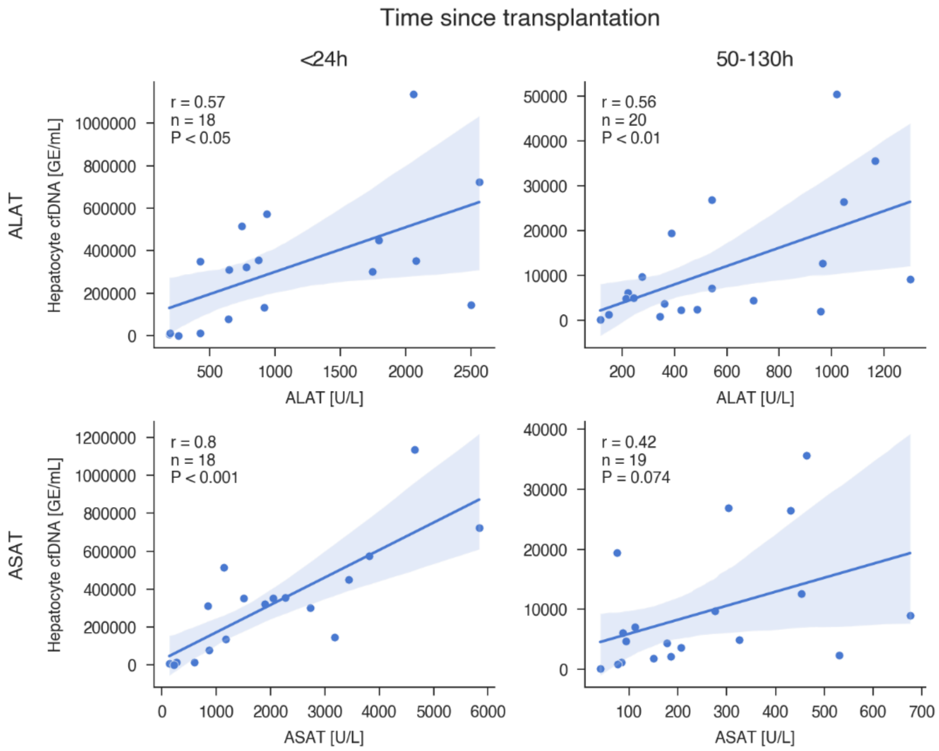


**Supplementary Figure 17: Correlation between absolute hepatocyte cfDNA and liver enzyme measurements in liver transplant recipients from the early post-transplantation cohort.**
The blue lines represent the linear regression with its corresponding 95%-confidence interval. Shown is the Pearson's r correlation coefficient.
